# Wastewater Based Epidemiology Enabled Surveillance of Antibiotic Resistance

**DOI:** 10.1101/2021.06.01.21258164

**Authors:** M.V. Riquelme, E. Garner, S. Gupta, J. Metch, N. Zhu, M.F. Blair, G. Arango-Argoty, A. Maile-Moskowitz, A. Li, C-F Flach, D.S. Aga, I. Nambi, D.G.J. Larsson, H. Bürgmann, T. Zhang, A. Pruden, P.J. Vikesland

## Abstract

Wastewater-based epidemiology (WBE) for disease monitoring is highly promising, but requires consistent methodologies that incorporate predetermined objectives, targets, and metrics. We demonstrate a comprehensive metagenomics-based approach for global surveillance of antibiotic resistance in sewage, enabling assessment of: 1) which antibiotic resistance genes (ARGs) are shared across regions/communities; 2) which ARGs are discriminatory; and 3) factors associated with overall trends including antibiotic concentrations in sewage. Across an internationally-sourced transect of sewage samples collected using a centralized, standardized protocol, ARG relative abundances (16S rRNA gene-normalized) were highest in Hong Kong and India and lowest in Sweden and Switzerland, reflecting national policy, measured antibiotic concentrations, and metal resistance genes. Asian versus European/US resistomes were distinct, with macrolide-lincosamide-streptogramin, phenicol, quinolone, and tetracycline versus multidrug resistance ARGs being discriminatory, respectively. Sales data were not predictive of antibiotics measured in sewage, emphasizing need for direct measurements. The WBE approach defined herein demonstrates multi-site comparability and sensitivity to local/regional factors.

---

Wastewater-based epidemiology (WBE) is a rapidly emerging framework for public health surveillance as it quickly and non-invasively provides anonymous population-scale information about human disease and anthropogenic chemical use.[1] Activity in this field has focused on xenobiotic and human biomarker measurements[2, 3] and the monitoring of infectious diseases such as polio[4] and typhoid.[5] Most recently, WBE has been proposed for surveillance of the spread of antibiotic resistance[6-8] as well as COVID-19.[9, 10] Within any given sewershed, the analysis of temporal or spatial changes in WBE targets (e.g., genes, microbes, chemicals) could provide an early warning of local and regional disease outbreaks,[11] while comparisons across sewersheds can enable insights into environmental and socioeconomic factors contributing to disease patterns.[12] To produce globally actionable information, however, data comparability across disparate systems is crucial and should be considered at the outset of commencing measurements. In this study, we applied a consistent sample collection and analysis protocol[13, 14] and demonstrated the capability of an array of data analysis approaches[15-17] for discriminating an international transect of sewage samples from Asia (Hong Kong, India, the Philippines), Europe (Sweden, Switzerland), and North America (US) with the aim of advancing a WBE framework for antibiotic resistance surveillance.

Sewage represents a composite of human-associated flora; including pathogens, antibiotic resistant bacteria (ARB), and antibiotic resistance genes (ARGs),[18-21] carried across the corresponding community and thus provides opportunities for environmental antibiotic resistance surveillance.[6, 22] The profiles of ARB and ARGs in sewage are expected to be dependent on current and historic antibiotic resistance management practices, including the types and quantities of antibiotics used and local attention to transmission control.[18, 23] A targeted and well-designed WBE program could help establish baseline levels of antibiotic resistance for a given community, inform effective antibiotic use policy and wastewater treatment practices, and serve as a point of comparison for assessing the impacts of such interventions.[12, 22]

Shotgun metagenomic sequencing provides a powerful means to characterize sewage microbiomes[24] and viromes,[25] producing nucleotide sequences that can be archived and compared to publicly-available databases to profile pathogens, pandemic viruses,[26, 27] and ARG composition (i.e., the “resistome”).[28-30] Recent studies have applied metagenomic sequencing to identify compelling sewage resistome trends. Hendriksen et al.[6] collected wastewater samples from 79 sites in 60 countries and found that ARG abundance correlated with socioeconomic, health, and environmental factors. Using the same dataset, Karkman et al. explored to what extent sewage metagenomes (alone or in combination with socioeconomic factors) predict local clinical resistance.[8] Pärnänen et al.[7] showed that sewage samples collected across a European North-South transect correlated with differences in antibiotic usage and average local temperatures. Going forward, precise surveillance objectives, targets, and metrics must be identified and validated if metagenomics enabled WBE is to be globally recommended and adopted.[22] In the context of antibiotic resistance, identification of informative features of resistomes and their relationship to other key sewage constituents, including concentrations of residual antibiotics, metals, metal resistance genes (MRGs), and mobile genetic elements (MGEs), is needed to gain insight to the factors driving resistance proliferation (**Figure 1**).

**Figure 1.**
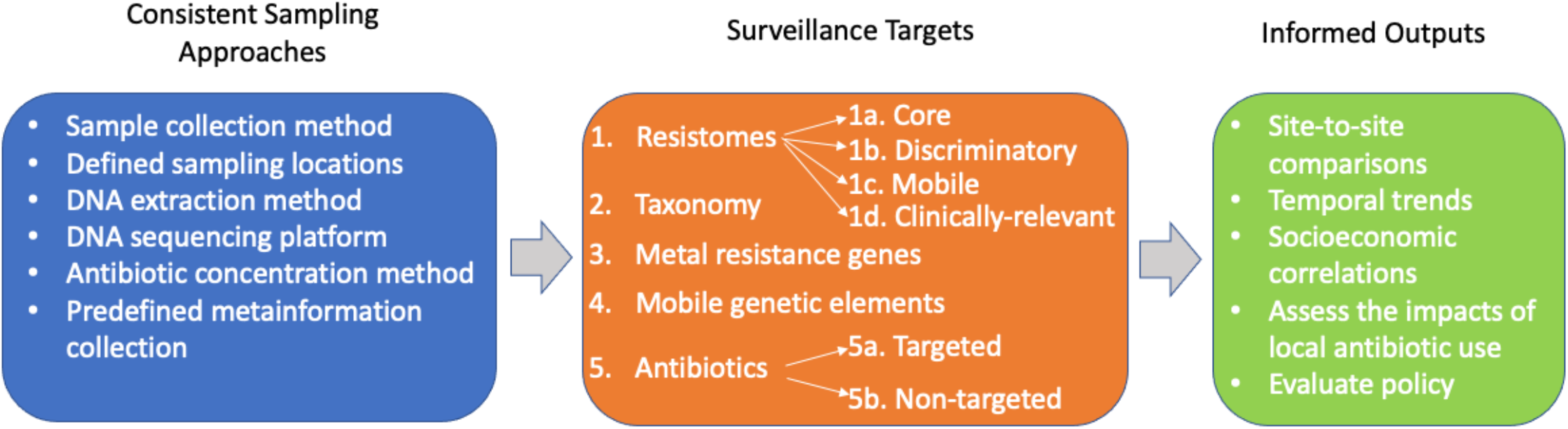
General strategy for antibiotic resistance monitoring.

Here we assessed various dimensions of sewage metagenomes, antibiotic concentrations, and the associated metainformation, to identify discriminatory features and factors that might reflect the corresponding resistomes. Taxonomic composition and the diversity of the corresponding microbiomes were evaluated as key constraints on ARG composition.[31, 32] In addition, plasmids, transposons, and integrons were quantified as plausible carriers of resistance[28, 29, 33] (i.e., indicating the potential for associated ARGs to spread horizontally across species). Genes that could provide co-selection opportunities, such as MRGs, were also quantified, addressing concerns that even if antibiotic use is curbed, metals and other selective agents could still select for ARGs. Finally, “core resistome”[31, 33, 34] and “discriminatory resistome”[16] analyses were carried out to account for the ARG “background” of globally-distributed or naturally-prevalent ARGs (e.g., those found even in 30,000 year old Arctic soils[35] and permafost[36]), and to differentiate ARGs that result primarily from anthropogenic activity and are of clinical concern. Herein, we advance a WBE framework for sewage-based surveillance of antibiotic resistance, focusing monitoring towards targets relevant to local evolution and dissemination.

## RESULTS AND DISCUSSION

### Comparison of Resistomes Across a Global Transect of Sewage Samples

Fourteen influent sewage samples from twelve WWTPs located in six countries were systematically collected by our team using standardized protocols,[13, 14] profiled, and compared. To identify and quantify ARGs, we performed shotgun metagenomic sequencing (20-30 million reads per sample; **Supplementary Table S1**) and annotated reads using the CARD database.[37] We detected an average of 449 ARGs (Range: 309-489) at each site, with the Chao index estimating true ARG richness ranging from 501-683 ARGs/sample (Average: 577; **Supplementary Figure S1**). When considering 16S rRNA gene normalized relative abundances, a clear trend emerged, whereby Asian sewage samples contained higher relative abundances of ARGs than European/US samples (**Figure 2A, Supplementary Figures S2** and **S3, Supplementary Data 1**). Such a pattern is likely driven primarily by the relative inputs of ARGs and ARBs from human populations, although other factors could be at play, including differences in relative industrial/hospital inputs or differential selection pressures when sewage travels through the collection network.[38, 39]

**Figure 2.**
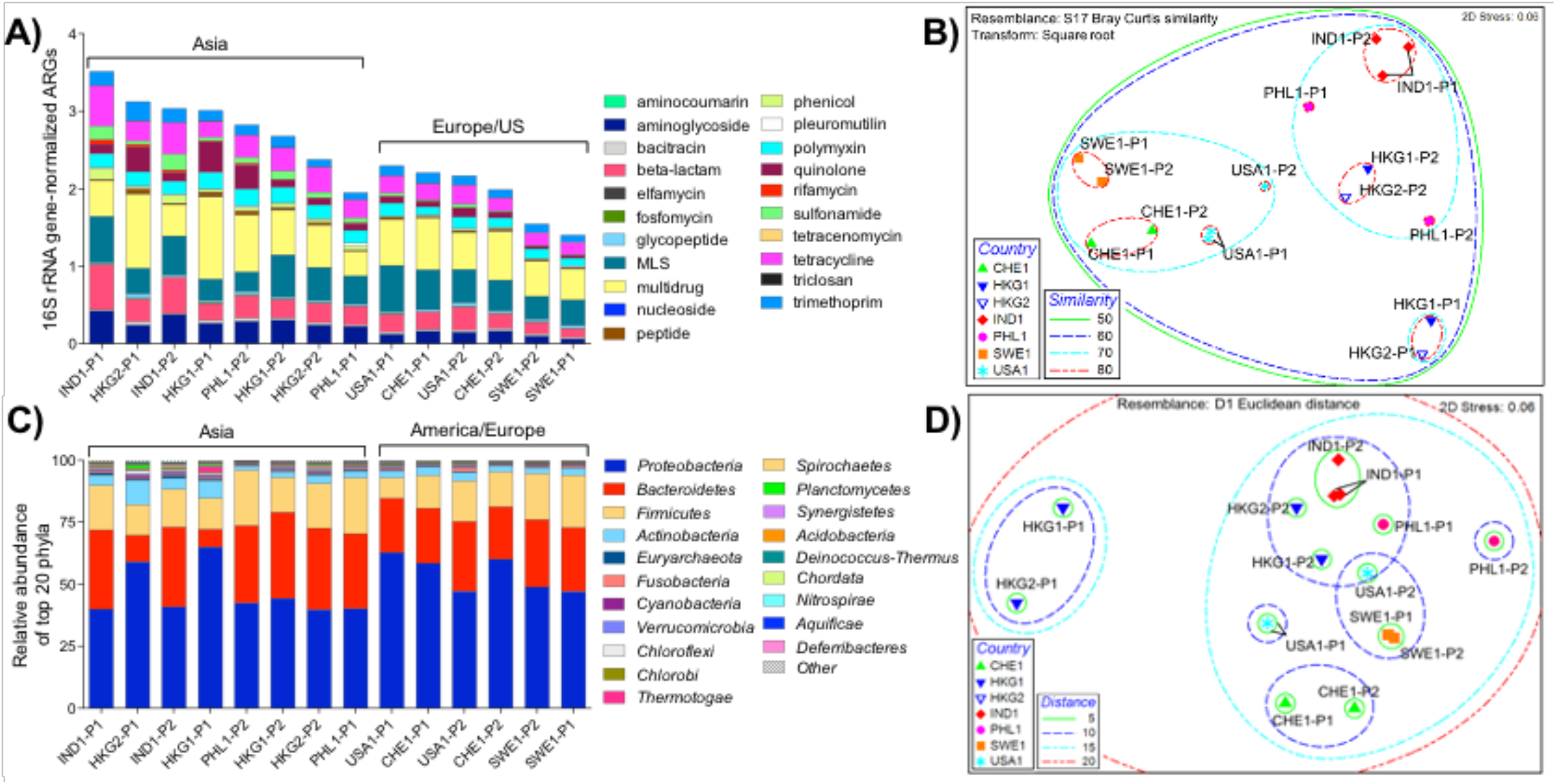
**A**: ARG distribution and relative abundance by corresponding antibiotic categories. ARGs were annotated via MetaStorm using CARD version 1.2.1. ARG categories were assigned in-house (**Supplementary Data 7**). Genes corresponding to two or more categories were labeled as “multidrug.” ARG abundances were normalized via MetaStorm to 16S rRNA gene abundances. **B:** Non-metric multidimensional scaling (NMDS) ordination of WWTPs according to ARG-based Bray-Curtis distance. Ellipses enclose sites of noted similarities, ranging from 0 to 100 (perfect similarity); thus, ellipses with similarity of 80, represent sites with highest similarity **C**: Relative abundance of top 20 bacterial phyla in WWTP influents. Genus-level annotations were done via MG-RAST using the Refseq database. **D:** NMDS ordination of WWTPs according to Euclidian distances between rlog-transformed reads to bacterial genera. Ellipses enclose sites of noted distances, where the shortest distance denotes the highest resemblance and longer distance denotes the least resemblance among the sites. Sample names refer to countries (IND: India, PHL: Philippines, USA: United States, CHE: Switzerland, HKG: Hong Kong, SWE: Sweden) along with the visit number (1,2) and WWTP plant number (P).

Hendriksen et al.[6] recently reported a global sewage survey and, although the analytical approach was distinct, the trend in ARG relative abundance was strikingly similar to the present study: higher in Africa (median: 2,100 fragments per kilo base per million fragments [FPKM]), Asia (1,200 FPKM), South America (1,900 FPKM), and the Middle East (1,100 FPKM), relative to Europe (750 FPKM), North America (900 FPKM), and Oceania (800 FPKM). Hendriksen et al.[6] applied FPKM normalization to address highly variable sequencing library depth (8-398 million reads per sample), while in the present study (20-30 million reads per sample) we normalized to 16S rRNA genes as a biomarker of total bacteria and correspondingly indicate the proportion of bacteria carrying ARGs.[40] Hendriksen et al. applied the ResFinder database[41] for ARG annotation, which resulted in detection of an apparently larger number of ARGs. This difference may result from the inclusion of multiple similar variants of the same ARG in ResFinder, but not in CARD, or from the inclusion of multiple ARGs that result from detection of a single ARG cluster. The differential number of ARGs included in different databases reflects the rapid evolution of the field of resistome bioinformatics[42] and the need to improve comparability across studies for sewage surveillance.[43]

We visualized ARG compositional similarities via non-metric multidimensional scaling (NMDS) ordination derived from Bray-Curtis dissimilarity matrices (**Figure 2B**). Clustering within the NMDS plot was tightest for WWTP influents originating from the same country, with the exception of the two Philippines WWTPs. A second level of clustering was observed between the Asian and European/US samples. One exception was the HKG-P1 influent, which clustered separately from all other samples. This latter result is consistent with the saline nature of this sewage, where ocean water is used for toilet flushing. Analysis of similarity (ANOSIM) indicated that sewage samples strongly separated from one another when grouped by location (R=0.741, p=0.001). Clusters also strongly separated when grouped as Asia versus Europe/US (R=0.700; p=0.002). Three clusters, less strongly separated (R=0.586; p=0.001), were apparent when grouped by continent. Importantly, two sets of temporally independent influent samples collected from two Hong Kong WWTPs (HKG1-P1/HKG2-P1 vs. HKG1-P2/HKG2-P2) clustered separately, but reproducibly, thus indicating less variation with time relative to geographic location. Ju et al.[31] also noted a high level of repeatability when characterizing resistomes in sewage samples from different WWTPs in Switzerland. Other studies[6, 44] have similarly shown locational reproducibility, which is key for inferring that observed differences are primarily driven by geographical location rather than the particular time of sample collection.

A number of clinically relevant ARGs (**Supplementary Data 2)** were detected in the influent of all WWTPs. OXA-type carbapenemase ARGs were ubiquitous and were detected at relatively high abundance (0.03-0.16 copies/16S rRNA gene copies). A variety of other β-lactamase ARG types (GES, CARB, SHV, CTX-M, and TEM) were also broadly detected, although at lower abundances. The *qnr*S gene (quinolone resistance) was detected in all sewages (0.001-0.24 copies/16S rRNA gene copies) and tended towards higher abundance in Asia (0.085±0.099 copies / 16S rRNA gene copies) relative to Europe/US (0.012±0.011 copies/16S rRNA gene copies), but not significantly so (Wilcox, p=0.1649). Others have noted differences in *qnr* carriage across populations,[45, 46] but statistical power here may have been insufficient to detect a difference. Reads annotated as the mcr*-1* colistin ARG were found at all WWTPs, with the exception of one Hong Kong plant.

### Core and Discriminatory Aspects of Resistomes

We subjected the collected data to “core” and “discriminatory” resistome analysis to further delineate similarities and differences between sites. ARGs and resistance classes that were ubiquitous across all sites reflect the “core”, while those that enable locational differentiation reflect “discriminatory” resistomes. The core resistome of 216 ARGs (**Supplementary Data 3**) was comprised of ARGs conferring multidrug resistance and resistance to aminoglycosides, β-lactams, macrolide-lincosamide-streptogramins (MLS), polymyxins, tetracyclines, and trimethoprims. The 25 most abundant ARGs found across all locations included one polymyxin (*pmr*E), one trimethoprim (*dfr*E), two tetracycline (*tet*Q and *tet*C), seven MLS (*msr*E, *mph*D, *erm*F, *mel, mac*B, *mef*A, *erm*B), one sulfonamide (*sul*1), three multidrug (*CRP, msb*A, *ade*J), two quinolone (*qnrS2, qac*H), one β-lactam (*cfx*A6) and two aminoglycoside (*aph(3’’)-lb* and *aph(6)-ld*) ARGs (**Supplementary Table S2**). Notably, among the core ARGs for which information was available, many were “ancient” or “natural background” ARGs identified in prior studies of pristine or less human-impacted environments (e.g., isolated caves, glacial soil, permafrost); thus, these ARGs likely predate the antibiotic era **(Supplementary Data 3)**. However, other core ARGs are known to have become globally distributed more recently (e.g., *sul*1, *sul*2, *dfrA*3, *tet*(G), *aad*A8). Based on pairwise comparison, the number of ARGs shared between sites ranged from 296 to 401 (**Supplementary Figure S4** with 349 average shared annotations. The overall proportion of core ARGs averaged 69% (min=62%, IND1-P2; max=72%, HKG2-P1). Notably, the least number of reads in common were between one of the Swedish (SWE1-P1) versus one of the Hong Kong sewages (HKG2-P1), which aligns with the Asia versus Europe/US divide in sewage resistome similarity. Consistent with the repeatability of the sampling events, the greatest number of common ARGs was noted for two Hong Kong sewage samples collected six months apart.

To focus analysis on the “variable” resistome, we removed core ARGs from the profiles in **Figure 2A**. This accentuated the continental differences in the sewage resistomes (**Figure 3**). Specifically, the relative proportions of aminoglycoside, β-lactam, rifamycin, sulfonamide, trimethoprim, and quinolone ARGs tended to increase, while those of MLS, multidrug, polymyxin, and tetracycline ARGs tended to decrease. Interestingly, the proportion of core ARGs tended to increase from Asia to Europe/US, which hypothetically could reflect a longer history of antibiotic use and management in Europe/US.

**Figure 3.**
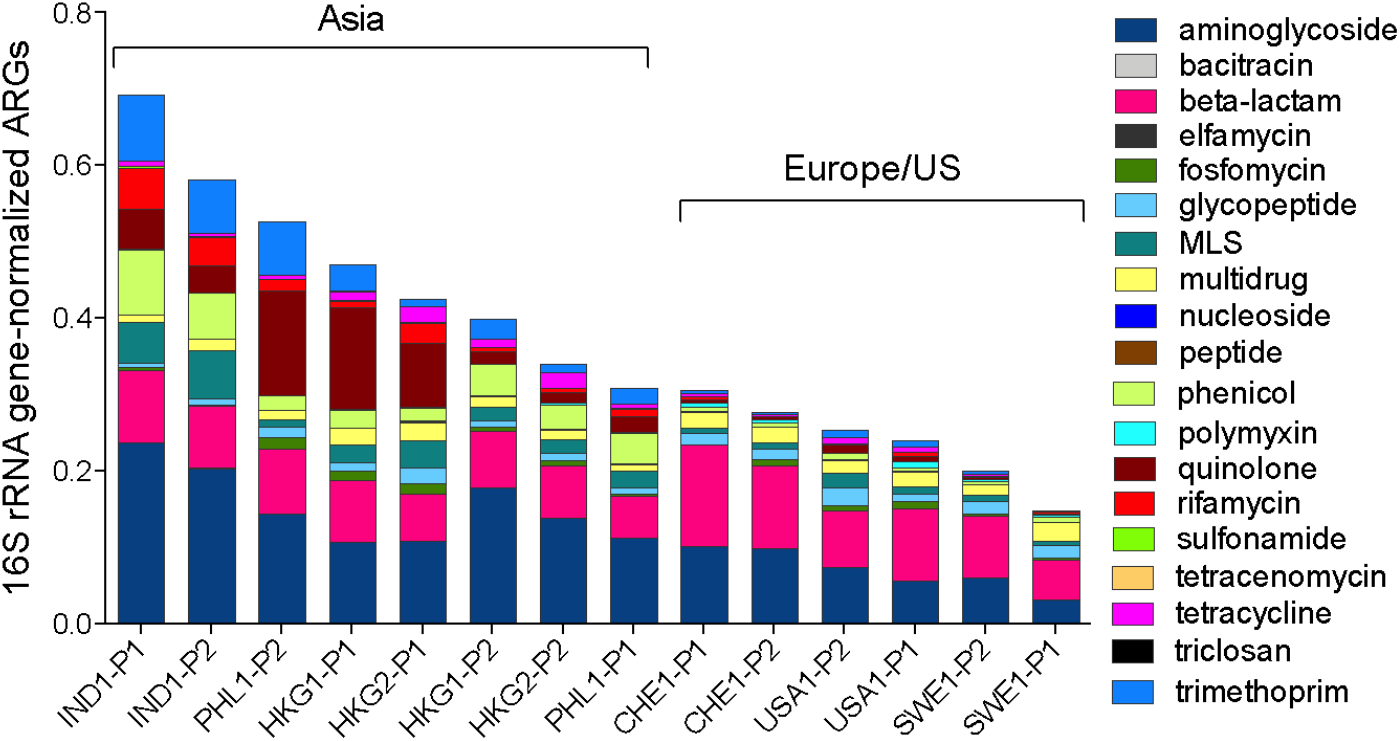
Distribution and relative abundance of ARGs comprising the “variable” resistome, i.e., with “core” ARGs removed from the analysis, by corresponding antibiotic categories.

We next characterized the “discriminatory” resistomes (i.e., ARGs that are, in terms of presence or abundance, indicative of local population or region). We applied the ExtrARG machine learning-based algorithm[16] to identify ARGs that discriminate sewage samples based on *a priori* selected groups. Sewage samples were grouped at the continent level (i.e., Asia vs. Europe/US) and then the relative abundances of the 50 most discriminatory ARGs were visualized using a heatmap (**Figure 4**). Two aminoglycoside ARGs, *aad*A8 and *aad*A2, were prominent in Asia, while ant*(3”)-IIb, aac(3)-Ia* and *axy*X were more prominent in Europe/US. Various β-lactam ARGs (*bla*_CphA5_, *bla*_OXA-363_, *bla*_OXA-309_, *bla*_OXA-371_, *bla*_FOX-8_, *bla*_FOX-1_, *bla*_CPS-1_) were abundant in Europe/US, but were found at low concentrations in Asian sewage. Interestingly, *bla*_CARB-2_ and *bla*_CARB-4_ were not detected in European/US sewage, but were found in Asia. The MLS ARG *erm*T, phenicol ARGs *cml*A1 and *cml*A4, quinolone ARG *qnr*VC6, and the tetracycline ARG *tet*(L) were primarily detected in Asian sewage. The sulfonamide ARG, *sul*2, was 3× to 5× more abundant in Asian sewage, while a number of the multidrug ARGs (e.g., efflux pumps *ade*J, *mex*K, *abe*M), were 3× to 15× higher in sewage from Europe/US. Further, we also found that a number of ARGs identified as discriminatory were only discovered within the last decade (e.g., ant*(3”)-IIb, axy*X, *bla*_CphA5,_ *bla*_CPS-1_, *bla*_FOX-8_, *qnr*VC6 and *qnr*VC6; **Supplementary Data 4**). We suggest that the location-specific distribution of discriminatory ARGs in sewage can be used to determine if their global distribution is widening. Such a result would suggest potential clinical concern.

**Figure 4.**
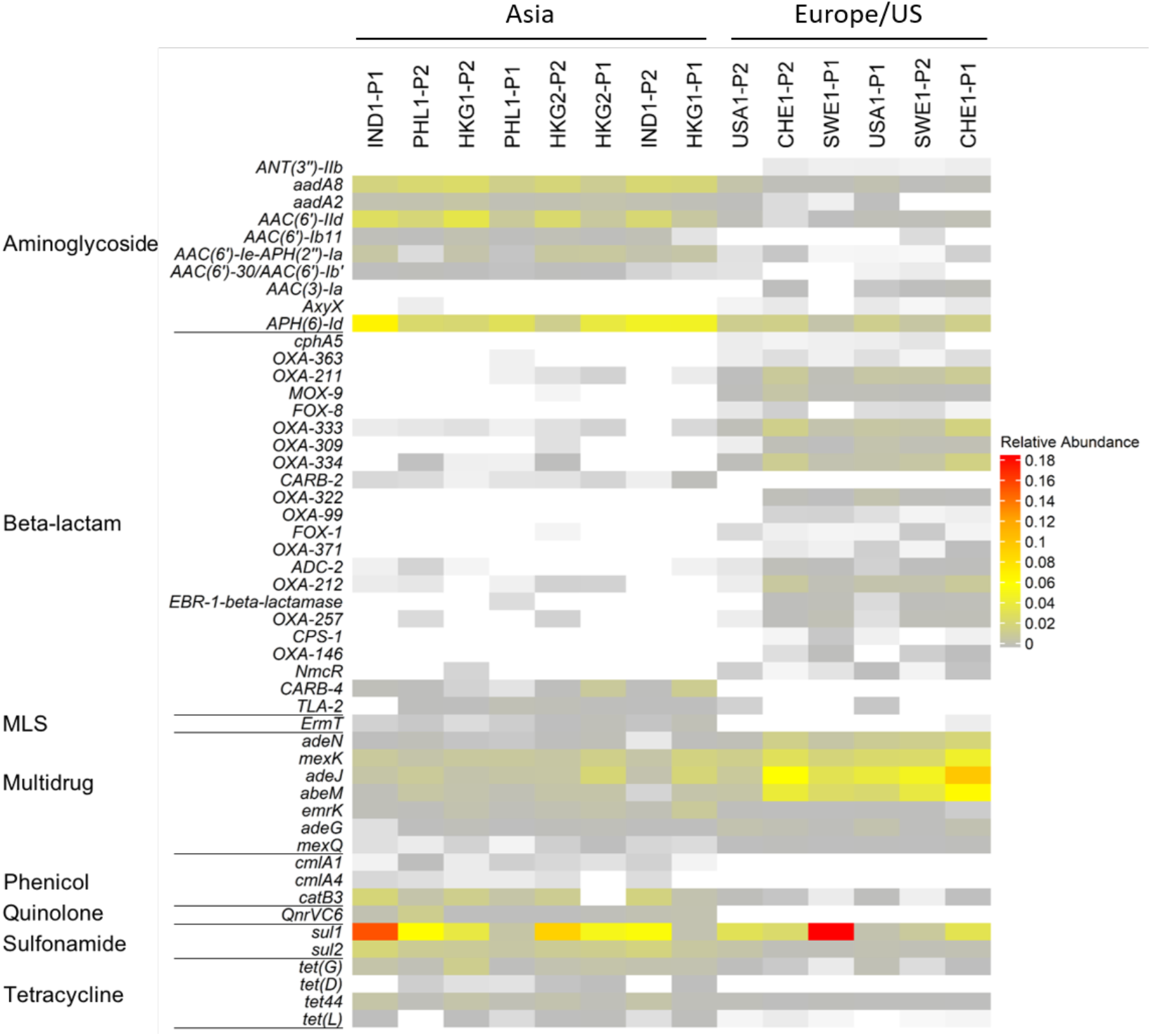
Heatmap representing the relative abundance of discriminatory ARGs for influent samples grouped according to their sampling continent identified using the ExtrARG python package.

### Comparison of Microbiomes Across the Global Transect of Sewage Samples

Similar to the resistomes, NMDS analysis of the sewage microbiomes (i.e., bacterial taxonomic composition derived from metagenomic data) resulted in clustering of samples as a function of geographic location (**Figure 2C** and **2D; Supplementary Figure S5**). ANOSIM indicated that the sewage microbiomes were most strongly separated when grouped by country (R=0.701, p=0.001). When grouped by Asia versus Europe/US, the groups were better separated (R=0.421, p=0.001) than when grouped by continent (i.e., Asia vs. Europe vs. US; R=0.3; p=0.003). As was the case for the resistomes, Hong Kong WWTP1 (HKG-P1) clustered separately from the other sewage samples. Similar patterns in the clustering of the resistomes and microbiomes are expected, since phylogenetic constraints to ARG carriage are well known.[31]

In accordance with the ARG-based class separations for Asia versus Europe/US, LEfSe analysis was performed on the microbiome data (**Supplementary Figure S6**). Four and ten classifying genera were identified in European/US versus Asian sewages, respectively. European/US sewages were distinguished by genera belonging to *Betaproteobacteria*, while *Clostridia* or *Negativicute* classes of the *Firmicute* phylum were discriminatory in Asian sewage. Only two non-*Firmicute*s genera (*Dehalococcoides* and *Shigella*) were found to be discriminatory in Asian sewage. Typically, higher levels of *Proteobacteria* are thought to reflect environmental conditions within the sewer system, while *Firmicutes* reflect greater human fecal influence.[47, 48]

Potential parallels between the microbiome and resistome compositions were investigated. First, the Mantel test was applied to either full or continent-specific distance matrices representative of the microbiome (Euclidian distances between log-transformed reads to bacterial genera) and resistome (Bray-Curtis dissimilarity matrix). When comparing all the collection sites, a strong positive correlation was observed between the ARG and genus-based taxonomy distance matrices (r = 0.74, p < 0.001). When separated by continent, a stronger correlation was observed between the Asian distance matrices (r = 0.80, p < 0.001), than between the European/USA distance matrices (r = 0.48, p = 0.03). For comparative purposes, a Bray-Curtis dissimilarity matrix was generated based on MetaPhlAn[49] output of taxonomic assignments obtained from MetaStorm[15] for Mantel-based comparison to the ARG similarity matrix. Again, a strong correlation was observed, both overall (r = 0.76, p < 0.001) and when compared continentally within Asia (r = 0.80, p < 0.001) and Europe/US (r = 0.56, p < 0.01).

Common pathogenic hosts of the discriminatory ARGs identified in **Figure 4** were also examined (**Supplementary Table S3**). Based on the data reported in the CARD database,[37] eighty-two pathogenic bacterial species are known to host these ARGs. Interestingly, discriminatory ARGs most representative of Asian sewages tended to have a broader range of potential host pathogens, whereas discriminatory ARGs associated with European/US sewages tended to have a narrower range.

### Genes Providing Opportunities for Co-Selection and Horizontal Gene Transfer

Genes documented to sometimes be found on plasmids were investigated as indicators of horizontal gene transfer potential. The average relative abundance of such genes identified via alignment to the ACLAME database was 56.7 copies/16S rRNA gene copies, with an observed minimum of 33.3 copies/16S rRNA gene copies in the Philippines (PHL1-P1) and a maximum of 111.6 copies/16S rRNA gene copies in Hong Kong (HKG2-P1; **Supplementary Figure S7**). No clear country- or continent-specific patterns were observed in terms of abundance.

Potential associations between ARGs and MRGs known to sometimes occur on plasmids or other MGEs were investigated and visualized via network analysis of assembled scaffolds (**Figure 5**). While acknowledging that there is uncertainty in the accuracy of scaffolds assembled from metagenomic data,[50] we carried out the analysis for empirical comparison assuming a consistent error rate across a sample set generated from the same sequencing platform. A substantial portion of reads were assembled, averaging 34% and ranging from 24% (HKG1-P1) to 50% (PHL1-P2). For each sewage sample, the number of generated scaffolds ranged from 64,576 (USA1-P1) to 243,356 (CHE1-P2), with an average length of 719 bp. All of the major classes of ARGs were detected on the scaffolds, with the most frequently observed belonging to the multidrug, MLS, glycopeptide, and tetracycline categories (**Supplementary Figure S8**). The relative distribution of ARGs (based on antibiotic category) among the scaffolds was remarkably similar for all sewage samples. Across all locations, the most common ARGs with plasmid associations were *pmr*E (peptide ARG), *acr*B (multidrug efflux), *dfr*E (trimethoprim ARG), *Mux*B (multidrug efflux), and *ros*B (peptide ARG) with 301, 186, 133, 125, and 101 co-occurrences, respectively. *pmrE* and *dfr*E were present at high abundance at all of the sites, thus suggesting globalization of these genes. Other notable plasmid ARG co-occurrences include *sul*1 (sulfonamide ARG, 20 co-occurrences), *MCR-*3 (peptide, 32 co-occurrences), and the *bla*_CARB,_ *bla*_OXA_, and *bla*_TEM_ β-lactam ARGs (between 1-13 co-occurrences). *sul*1 is known to be highly associated with both class 1 integrons[51] as well as plasmids. Some of these co-occurrences correspond to resistance against carbapenems and colistin, which are critically-important antibiotics of last resort. This analysis illustrates that clinically-important classes of ARGs were readily found on assembled scaffolds that also contained genes commonly found on plasmids, an indicator that they are potentially in a mobile form in sewage.

**Figure 5.**
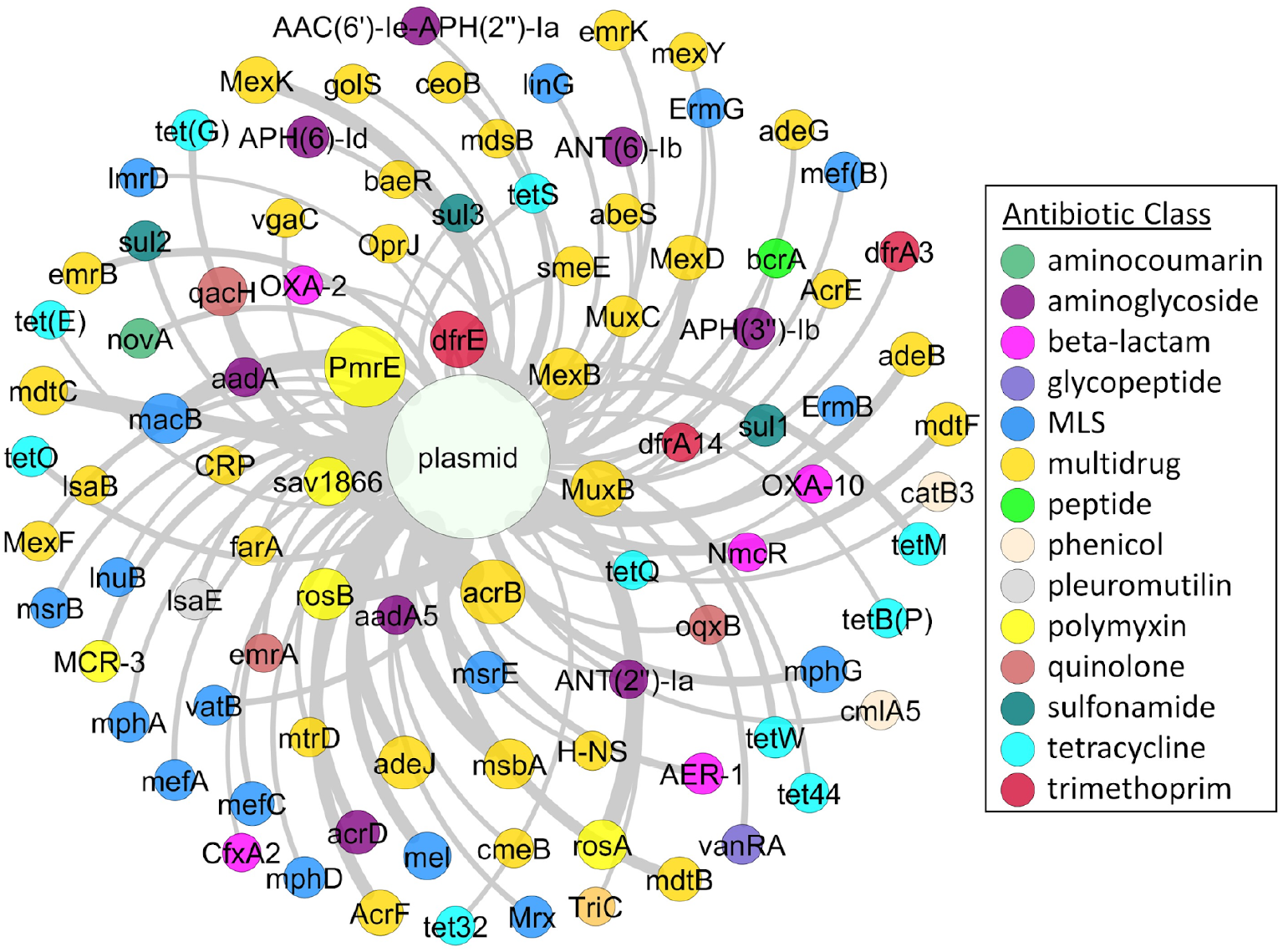
Co-occurrence of ARGs and plasmid gene markers on *de novo* assembled scaffolds generated by shotgun metagenomic sequencing reads pooled from all samples. This analysis highlights ARGs that are probable candidates for potential horizontal gene transfer (co-occurrences with plasmid associated genes) or co-selection (co-occurrences with ARGs of different classes). Proximity of nodes and width of lines indicate frequency of associations between genes. Node diameter is proportional to the number of co-occurrences for that gene. Co-occurences with fewer than 3 instances were excluded from the network analysis rendering. MLS = macrolide, lincosamide, and streptogramin resistance.

ARGs and MRGs are often subject to co-selection pressure when they are both present on a single genetic element, such as a plasmid, or cross-selection if the same gene is both an ARG and an MRG, as is commonly the case for multidrug efflux pumps capable of pumping both metals and antibiotics.[52] The ranking of total MRG relative abundances, identified via alignment to the BacMet database,[53] was strikingly similar to that of the total ARG relative abundances (Spearman r = 0.66, p = 0.013), except both Indian WWTPs and one Philippines plant ranked much lower than the other Asian WWTPs, while a Swiss WWTP had extremely low MRG relative abundance (**Figure 6**). This finding is suggestive that there are, or have been, common drivers, such as metals, antibiotics or other agents, either within human gastrointestinal tracts, in industrial inputs, or in sewers themselves, that exert (or have exerted) selection pressure for carriage of both ARGs and MRGs.

**Figure 6.**
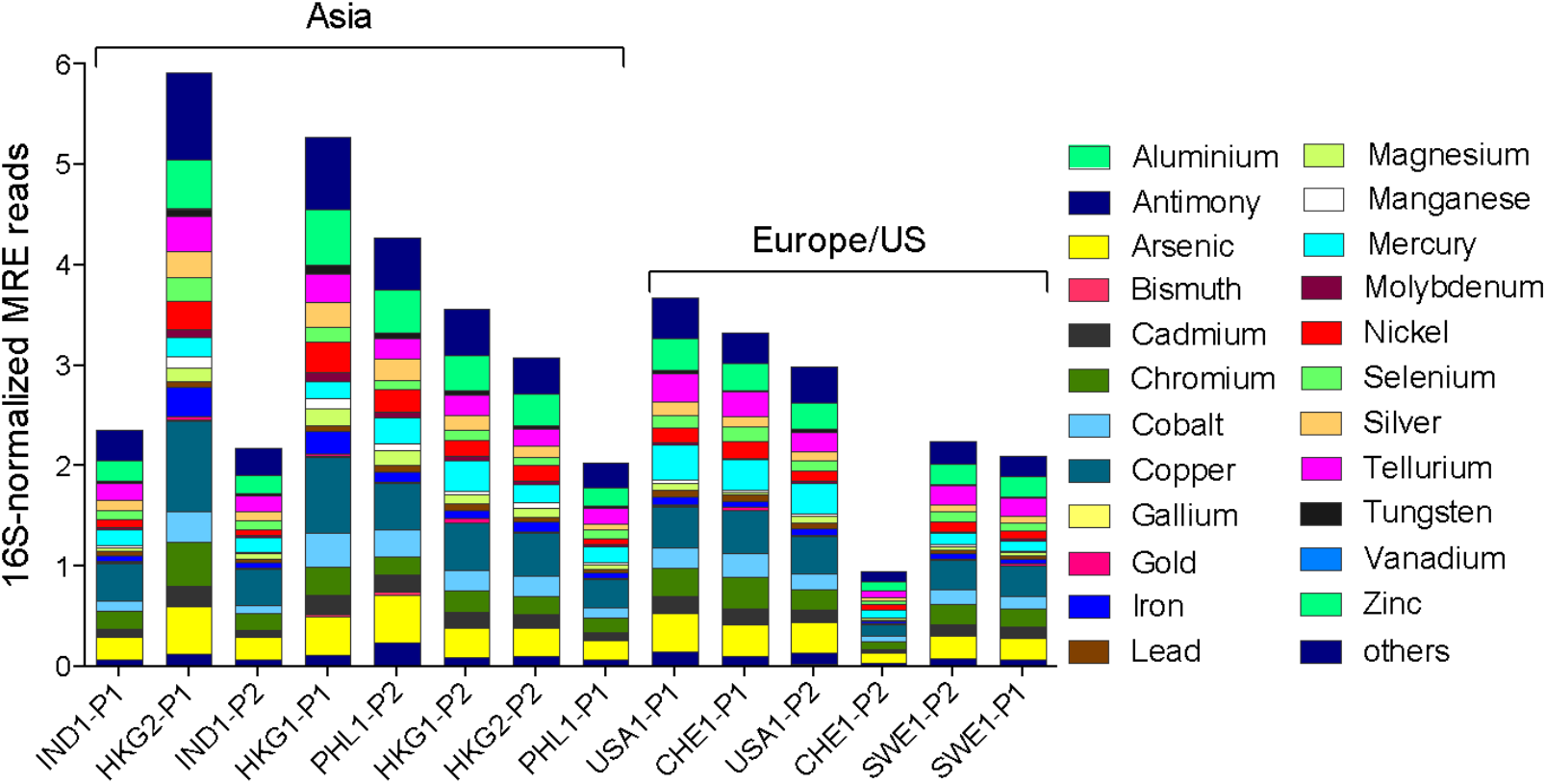
Distribution and relative abundance of metal resistance genes in WWTP influents. Metal resistance genes were annotated via MetaStorm using the BacMet database version 1.1. Meta resistance gene abundances were normalized to 16S rRNA gene abundances. Sample names refer to countries (IND: India, PHL: Philippines, USA: United States, CHE: Switzerland, HKG: Hong Kong, SWE: Sweden) along with the visit number and WWTP plant (P) number. Order is according to the ranked comparison of total ARG relative abundance shown in Figure 1.

Across all locations, multidrug ARGs were the most common type associated with MRGs (**Supplementary Figure S9**). The most common co-occurrences within the assembled scaffolds were noted between *acr*B and *Mex*B ARGs, co-located with copper/zinc MRGs (160 and 83 co-occurrences, respectively), and *mux*B ARG co-located with zinc MRGs (96 co-occurrences). These co-locations were consistently among the most abundant at all sample sites. A literature review was conducted to examine whether any of these presumed co-occurrences were actually the same gene, and thus indicative of cross-rather than co-selection. We identified nine genes that were consistently abundant across all the sample sites and confer resistance to both antibiotics and metals, namely, *mdt*A, *mdt*B, *mdt*C, *bae*S, *bae*R, *cme*B, *acr*D, *mex*I and *pmr*C. As anticipated, most of these genes are known to be associated with efflux pump systems (**Supplementary Data 5**).

### Sewage Antibiotic Concentrations

Antibiotic, pharmaceutical, and personal care product (PPCP) concentrations measured in these sewage samples were recently reported in detail by Singh et al.[14] and trends relevant to the present study are summarized in **Table 1** and **Supplementary Figure S10**. We note that chemical stability issues precluded quantification of the β-lactams and that, due to logistical constraints, the antibiotic measurements for the Indian sewage were made during a sampling trip that occurred one year later. We measured the highest antibiotic concentrations in Hong Kong sewage, where total concentrations of macrolides, quinolones, sulfonamides, tetracyclines, and trimethoprim reached levels >63,000 ng/L. The antibiotics with the highest concentrations in the sewage were ciprofloxacin (48,100 ng/L, HKG1-P1) and clarithromycin (39,551 ng/L, IND2-P2). In particular, Indian sewage contained very high levels of anhydro-erythromycin and norfloxacin. To contextualize these measured antibiotic concentrations, we first compared them to predicted no effect concentrations (PNECs; **Table 1**)[54] for resistance selection. All samples, except one from Sweden, were above the PNEC for ciprofloxacin, while norfloxacin was near or above the PNEC in both Swiss sewages and one Indian sewage sample. Clarithromycin was above the PNEC in one US sample. Two Hong Kong sewage samples had concentrations greater than the PNEC for the tetracyclines, while both the Hong Kong and Indian sewages contained the macrolides azithromycin and clarithromycin at levels exceeding the PNEC. It is noted that PNEC values provide a point of reference, but measured concentrations that either exceed or fall short of a PNEC do not necessarily delineate the presence or absence of selection pressure within sewage.[54] Resistance metrics quantified in sewage likely reflect both current and historic drivers, including past patterns of antibiotic use and selection pressures that may have occurred in the human gut, that collectively shape the resistome; however, antibiotic measurements can only reflect usage at the time of sampling.

**Table 1:** Proposed no effect concentrations (PNEC),^52^ representing the cutoff below which no selection of resistant bacteria is anticipated to occur and concentrations (ng/L) of antibiotics detected in the influent of each WWTP. Concentration data originally published in Singh et al. ^**13**^

|  |  | Asia |  |  |  |  |  |  |  | Europe/US |  |  |  |  |  |
| --- | --- | --- | --- | --- | --- | --- | --- | --- | --- | --- | --- | --- | --- | --- | --- |
| PNEC (ng/L) | COMPOUND | HKG1-P1 | HKG1-P2 | HKG2-P1 | HKG2-P2 | IND2-P1* | IND2-P2* | PHL1-P1 | PHL1-P2 | SWE1-P1 | SWE1-P2 | CHE1-P1 | CHE1-P2 | USA1-P1 | USA1-P2 |
|  | Caffeine | 14846.1 | 11969.6 |  |  | 60880.6 | 22752.9 |  |  | 26884.8 | 35133 | 41781.1 | 6726.8 | 23673 | 16512 |
|  | Carbamazepine | 1576.3 | 391.4 | 309.7 | 100.2 | 270.6 | 88.6 | n.d. | 2.2 | 8176.8 | 7961 | 22.7 | 91.4 | 174.6 | 82.2 |
|  | Iopromide | 149.7 | 162.9 |  |  |  |  |  |  |  |  |  |  | 6401.4 | 2324 |
| 1000 | Tetracyclines | -- | -- | 11694.9 | 4790.6 | -- | 240.5 | 30.5 | n.d. | 170.3 | -- | -- | -- | 16.2 | 1.1 |
| 500 | Trimethoprim | 31.7 | 12 | 120.3 | 45.1 | -- | -- | < LOD | 50.5 | 38.5 | 39.1 | -- | 121.3 | 438.6 | 137.2 |
|  | Acetylsulfamethoxazole | 338 | 97.2 | -- | -- | -- | 377.1 | 394.9 | 195.4 | 145.3 | 38.1 | 296.8 | 419.5 | 1214.2 | 634.8 |
|  | Sulfadiazine | -- | -- | -- | -- | -- | -- | -- | -- | -- | -- | -- | -- | -- | -- |
|  | Sulfamethoxydiazine | 66.4 | 36.5 | -- | -- | -- | -- | -- | -- | -- | -- | -- | -- | -- | -- |
| 16000 | Sulfamethoxazole | 124.6 | 107 | -- | -- | -- | -- | 360.4 | -- | 21 | 5.7 | 175.5 | 177.6 | -- | 1941.3 |
|  | **ΣSulfonamides** | 529 | 240.7 | -- | -- | -- | 377.1 | 755.3 | 195.4 | 166.3 | 43.9 | 472.3 | 597.1 | 1214.2 | 2576.2 |
|  | **ΣSulfonamides+Trimethoprim** | 560.7 | 252.7 |  |  |  | 377.1 | 755.3 | 245.9 | 204.8 | 83 | 472.3 | 718.4 | 1652.8 | 2713.4 |
| 1000 | Anhydro-Erythromycin | -- | -- | 222 | 86.2 | 310 | 1894 | 467.3 | 230.4 | 126.2 | 19 | 1.5 | 18.5 | -- | 55.8 |
| 1000 | Roxithromycin | 263.1 | 85.2 | -- | -- | -- | -- | -- | -- | -- | -- | -- | -- | -- | -- |
| 250 | Azithromycin | 4585.1 | -- | 1213.2 | 249.5 | 239.4 | 4127.5 | -- | -- | n.d. | 17.1 | 14.2 | 41.7 | -- | 212.2 |
| 250 | Clarithromycin | 10107.6 | 1282.1 | 1473 | 490.7 | 65.8 | 39551.3 | 97.2 | 58.6 | 8.3 | 5 | 18.6 | 206.6 | -- | 387.3 |
|  | **ΣMacrolides** | 14955.7 | 1367.3 | 2908.2 | 826.4 | 615.1 | 45572.8 | 564.5 | 289 | 134.6 | 41 | 34.2 | 266.8 | -- | 655.2 |
| 64 | Ciprofloxacin | 48103.1 | 146 | 1620.9 | 521.6 | 1589.1 | 1357.6 | 567.4 | 375.1 | 58 | 86.2 | 825.5 | 2505.3 | 1090.2 | 514.8 |
| 64 | Enrofloxacin | -- | -- | -- | -- | 45.6 | -- | -- | -- | -- | -- | -- | -- | -- | -- |
| 500 | Norfloxacin | 75.7 | 8 | 155.6 | 45.6 | 1853.8 | 430.1 | -- | -- | -- | -- | 438.8 | 803.2 | -- | -- |
|  | **ΣFluoroquinolones** | 48178.8 | 154 | 1776.6 | 567.2 | 3488.5 | 1787.8 | 567.4 | 375.1 | 58 | 86.2 | 1264.4 | 3308.5 | 1090.2 | 514.8 |

Note: Highlighted rows reflect total concentrations of the given class of antibiotic.
\*Antibiotic measurements were made for Indian sewage samples collected one year after the samples subject to metagenomic analysis

To further contextualize the measured antibiotic concentrations, we compared them to values predicted based upon antibiotic sales data. Prior studies have used sales data to explain global differences in resistomes in sewage[6] and other contexts.[55] Antibiotic consumption data between 2000-2015 for each site was obtained from ResistanceMap.^[56]^ As shown in **Supplementary Figure S11**, total antibiotic consumption (defined daily doses (DDD) per 1000 population) was reported to be highest in the US, followed by Hong Kong, Sweden, and Switzerland.[55] Reported antibiotic use in India and the Philippines has historically been lower than that in Europe/US, but in recent years, reported use in India has increased and is estimated to be similar to that in Sweden and Switzerland as of the 2015. Reported antibiotic use in the Philippines lags relative to other countries and has remained fairly stable over the 2000-2015 period.

We calculated *expected* antibiotic concentrations within our sewage samples based upon sales data, data on pharmaceutical excretion rates, and published relationships between xenobiotic analyte concentrations and sewershed populations. As illustrated in **Supplementary Figure S10**, these estimated antibiotic concentrations overestimate the measured antibiotic concentrations by orders of magnitude. Such overestimates hold true regardless of the locality or the xenobiotic used. Similar overestimates were also found using estimated sewershed populations and sewage flow rates employed. These disparities are unsurprising given the many potential disconnects at the national, local, and regional level between antibiotic sales, consumption, and disposal patterns[57, 58] as well as the wide differences in biological and chemical decay rates exhibited by these antibiotics.[59] Considering that measured antibiotic data better reflected measured trends in the resistome, (i.e., Hong Kong and Indian sewage ranked highest in total ARG relative abundance; **Figure 2A**), we recommend that determination of actual antibiotic concentrations in sewage should take precedence over extrapolations from general sales or consumption reports. Because antibiotic use often follows seasonal patterns, ideally such measurements should take place regularly throughout the year. It is recognized, however, that some antibiotics (e.g., beta lactams) have short half-lives and/or recoveries and thus, while better, direct measurement of antibiotics in sewage is not a perfect solution.

We also explored if prior reports of links between antibiotic concentrations and resistant clinical isolates^58,59^ held true for sewage resistomes. Bonferroni-corrected Spearman rank order correlation analyses were conducted to identify potential relationships between concentrations of antibiotics or pharmaceuticals and PPCPs, individual ARGs, and ARG classes (**Supplementary Data 6**). At the individual ARG level, few correlations were observed when comparing against antibiotic compound concentrations: clarithromycin concentration showed a positive correlation with *tet*(L) (≥ 0.93, p<0.01) and a negative correlation with *mph*B (≥ -0.91, p<0.05); and norfloxacin showed a positive correlation with the *cat* gene (≥ 0.91, p<0.05). At the ARG class level, macrolide antibiotics positively correlated with quinolone ARGs (≥ 0.84, p<0.05). Of the four macrolide antibiotics tested, clarithromycin presented the strongest correlations with quinolone ARGs. A remarkably similar pattern of macrolide antibiotics and quinolone ARG correlation was previously observed in Swiss sewage.^30^ Overall, few correlations between ARGs and antibiotics were observed and those that were observed pertained to macrolides. Lack of simplistic correlations between antibiotics and ARGs is consistent with prior work[6, 19] and is to be expected considering that the subset of antibiotics analyzed do not capture the full range of antibiotics actually present, while certain dominant ARGs may not be subject to selection pressure and others may be negatively selected by certain antibiotics. Also, effective concentrations of antibiotics may be too low to exert measurable selection pressure in the sewage or selection pressure might have occurred historically in the human gut and thus there is a spatiotemporal disconnect between the two measures.

### Surveillance Effectively Reveals Key Sewage Resistome Trends

We observed that the relative abundances of total ARGs were elevated in Asian versus European/US sewages, consistent with general perceptions of rigor in antibiotic use policy across countries[19] and the generally higher antibiotic loadings in Asia. The lowest relative abundances of ARGs were found in the sewage from Sweden which has incorporated numerous proactive measures to avoid misuse and overuse of antibiotics.[60] Hong Kong, India, and the Philippines, on the other hand, are characterized by dense urban populations and either have problems with illegal antibiotic sales without a prescription[61] or do not require prescriptions for antibiotic use.[57]

Removal of core ARGs (i.e., those ubiquitously detected in urban sewage) from the resistome profiles accentuated the differences between Asia and Europe/US, while delineation of discriminatory ARGs identified those that uniquely circulate in each corresponding region. In general, a greater variety of ARGs across multiple resistance classes were identified as discriminatory in Asian sewage, while a subset of aminoglycoside and multi-drug ARGs were characteristic of European/US sewages. Such knowledge provides information about the ARGs within local regions in terms of their prevalence, novelty, or their potential carriage by pathogens. Surveillance is a key component of the global action plan to combat antimicrobial resistance.[62] Within that context, consistent methods and approaches are required to facilitate robust data comparability (**Figure 1**). The approach outlined herein provides such consistency, but it would costly to apply everywhere due to its reliance on a dedicated sampling and analysis team. The alternative approach,[6] relying upon centralized processing of large numbers of shipped samples, has the capacity to more broadly elucidate global trends, while more focused efforts, such as that delineated herein, provide the potential for greater granularity (e.g., comparing local sewersheds, sampling within a sewershed). Implementation of coordinated sewage resistome surveillance efforts will be key to improved understanding of baseline resistome characteristics.[42] Such information can be used as a reference point for future sampling to aid in early identification of previously unobserved ARGs that may pose clinical threats, including the emergence of new resistance types.[22]

### Implications for Wastewater-Based Epidemiology

WBE is rapidly gaining attention as a highly promising tool for infectious disease monitoring.[2, 6, 7, 12, 33, 63] Here we demonstrated the potential for metagenomic-based surveillance of ARGs and other key metainformation in sewage to provide a foundation for WBE of antibiotic resistance, and we have illustrated challenges that must be considered as the field develops. In particular, it is critical to define WBE surveillance objectives; including what will be monitored, how samples (and appropriate metainformation) will be collected and analyzed, and how the data will ultimately be interrogated prior to investing in extensive sampling campaigns. The hypothesis that the sewage resistome composition differs by geographical location could only be supported through the use of consistent experimental and data analysis protocols, which will be particularly vital in future efforts to further dissect the role of geographical sub-factors, such as socioeconomics, population density, antibiotic use, diet, sanitation status, and local climate/temperature.[19, 20] Comparison of results obtained using widely varying methods (e.g., nucleic acid extraction kits,[13] solid-phase extraction protocols[14]) or metagenomic platforms present inherent and substantial limitations to identifying key factors driving the spread of antibiotic resistance.

For WBE to be applied towards estimating the prevalence of infectious disease or antibiotic resistance for a given region, reasonably accurate estimates of the population contributing to a sewage sample are needed.[2, 64] We utilized xenobiotic analyte measurements to develop estimates of the population within a given sewershed. These measurements enabled estimation of the *de facto*[64] population (i.e., incorporating transient contributors to the sewershed) at the time of sampling. A required input to these calculations is the sewage flow rate. Such metainformation is not necessarily collected in all studies and the lack of such information for the sewage samples from the Philippines precluded estimation of antibiotic concentrations for that country. An additional consideration of such population estimates is the assumption that relationships developed within Australia relating PPCP concentrations to population are globally applicable. Given disparities in antibiotic and PPCP use across the world this latter assumption requires evaluation. Incorporation of antibiotic and PPCP measurements within the context of WBE requires *a priori* planning if such measurements are to be used to estimate study populations within the sewershed. Given these considerations, alternative approaches to estimate human contributions to sewage (e.g., quantification of pepper mottle mild virus[65] or crAssphage[21]) may be preferable.

The WBE approach defined herein demonstrates a means to identify geographic “hot-spots” where attention towards curbing the spread of antibiotic resistance may be particularly warranted. This is especially relevant as we emerge from the new era of pandemic awareness and concern, where multidrug resistant pathogens, ARGs, and infectious viruses have been observed to rapidly spread across international borders.[66] Estimates that several thousands of deaths per year globally can be attributed to antibiotic resistance amplifies the need for coordinated, global surveillance. Ultimately, widescale online implementation of metagenomic surveillance of ARGs and other disease determinants within sewage in the context of a WBE framework promises to yield new insights. Such insights will help illuminate the specific factors that drive the dissemination and spread not only of antibiotic resistance, but also other diseases of emerging concern.

## METHODS

### Sample locations

A total of 14 sewage samples were collected from 12 WWTPs located in India, Hong Kong, the Philippines, Sweden, Switzerland and the US, with 2 WWTPs per location and the Hong Kong WWTPs subjected to two separate sampling events. The WWTP capacities ranged from 2.6-66 MGD with a majority of the sewage consisting of domestic wastewater, with some industrial and hospital inputs (**Table 2**).

**Table 2:** Sewage sample characteristics

| Country | Sample ID | Sampling Date | WWTP<br>Capacity (MGD) | Sewage Composition | Temperature<br>(°C) | Dissolved Oxygen<br>(mg/L) | pH |
| --- | --- | --- | --- | --- | --- | --- | --- |
| **Asian** |  |  |  |  |  |  |  |
| Hong Kong | HKG1-P1 | 14.07.2016 | 66 | Municipal sewage | 31.8 | 0.51 | 7.80 |
|  | HKG1-P2 | 14.07.2016 | 22 | Municipal sewage +<br>treated seawater | 30.3 | 1.78 | 5.78 |
|  | HKG2-P1 | 06.12.2016 | 66 | Municipal sewage | 24.5 | 1.89 | 7.50 |
|  | HKG2-P2 | 06.12.2016 | 22 | Municipal sewage +<br>treated seawater | 25.6 | 1.61 | 7.30 |
| India | IND1-P1 | 10.03.2016 | 14 | **Municipal sewage** | 29.8 | 0.94 | 7.96 |
|  | IND1-P2 | 10.03.2016 | 14 | **Municipal sewage** | 30.9 | 0.22 | 7.64 |
| Philippines | PHL1-P1 | 02.12.2016 |  | Municipal sewage +<br>industrial input | 28.1 | 2.88 | 8.16 |
|  | PHL1-P2 | 29.11.2016 |  | Municipal sewage | 26.5 |  | 7.17 |
| **Europe/US** |  |  |  |  |  |  |  |
| Sweden | SWE1-P1 | 08.06.2016 | 26 | Municipal sewage +<br>hospital and industrial<br>input | 15 |  |  |
|  | SWE1-P2 | 09.06.2016 | 2.6 | Municipal sewage +<br>hospital and industrial<br>input | 16.5 |  |  |
| Switzerland | CHE1-P1 | 17.05.2016 | 32.6 | 90% municipal, 10%<br>industrial | 13.4 | 6.63 | 7.55 |
|  | CHE1-P2 | 18.05.2016 | 5.8 | 50% municipal, 50%<br>industrial | 16.4 | 4.22 | 8.17 |
| United States | USA1-P1 | 01.11.2016 | 6 | 95% municipal | 18.9 | 3.24 | 6.79 |
|  | USA1-P2 | 19.01.2017 | 13.45 | 80% municipal, 20%<br>industrial | 15.8 | 0.42 | 6.95 |

### Sample collection and processing

Sample collection and processing were standardized as described previously.[13, 14] In brief, influent grab samples were collected in sterile polypropylene containers and transported to a local laboratory on ice. Sewage biomass was concentrated in triplicate via vacuum filtration of equal influent volumes onto three separate 0.22 µm mixed cellulose ester membranes (MilliporeSigma Darmstadt, Germany), which were then preserved in 50% ethanol and stored at -20 °C. Ethanol-fixed membrane samples were shipped to Virginia Tech for DNA extraction and biomolecular analyses. Sewage samples (0.5 L) for chemical analysis were acidified and filtered using 0.45 µm glass microfiber filters to remove microorganisms and particulate matter. Na_2_EDTA (2 mL, 5% v/v in water) and isotopically labeled surrogate standards (50 µL of 1000 µg/L surrogate mix solution) were added to each sample.

### DNA extraction, shotgun metagenomic sequencing and analyses

As described previously,[13] DNA extraction was carried out using the FastDNA Spin Kit for Soil (MP Biomedicals, Solon, OH). Composite samples were prepared by pooling equal masses of triplicate samples. TrueSeq libraries (Illumina, San Diego, CA) were prepared for shotgun metagenomic sequencing via Illumina HiSeq 2500 with 2 × 100 paired-end reads. Sequencing was performed at the Virginia Tech Biocomplexity Institute Genomic Sequencing Center (Blacksburg, VA). Library preparation and sequencing were duplicated independently for two samples to verify sequencing reproducibility. After quality trimming, sequencing depth ranged from 10 to 16 million paired-end reads per sample with an average of 13 million reads per sample (**Supplementary Table S1**).

### Taxonomic, ARG, plasmid, and metal resistance gene (MRG) analyses

Sequences were uploaded to the metagenomics RAST server (MG-RAST) and annotated against the RefSeq database to identify bacterial phyla and genera using default parameters.^[67]^ Taxonomic beta diversity was examined via NMDS based ordination according to Euclidian distances between rlog-transformed reads to bacterial genera.[68] ARG annotation was carried out via MetaStorm[15] with read matching to CARD (protein homolog model) version 1.2.1.[69] The pipeline uses the DIAMOND BLASTX^71^ aligner with the representative hit approach (E-value<1e-10, identity>90%, and minimum length of 25 amino acids). The ARGs in CARD version 1.2.1 were assigned into resistance categories based on the updated ARGminer database.^[17]^ Manual curation was subsequently performed to fill in missing categories or to update existing categories and to segregate the genes thought to confer resistance via point mutations. Gene names and the ARG category to which they were assigned are found in **Supplementary Data 7**. If an ARG conferred resistance to two or more antibiotics classes, it was categorized as “multidrug.” The core resistome is defined as the collection of ARGs that are ubiquitous and are an inherent part of the resistome. Core resistome analysis was carried out by subsetting the list of ARGs that were identified in all samples (**Supplementary Data 3**). Plasmid-associated sequences were annotated using the ACLAME database, version 0.4.[70] MRGs were annotated using the list of experimentally confirmed metal resistance genes from BacMet, antibacterial biocide and metal resistance genes database, version 1.1.[53] Gene abundances were normalized within MetaStorm to 16S rRNA gene abundances as previously described.^[40]^ NMDS and analysis of similarities (ANOSIM) was conducted considering each individual ARG detected (i.e., not grouped by class) with square root transformation and Bray-Curtis dissimilarities. R-value cutoffs as defined by Clarke and Warwick[71] were used (R>0.75, well separated; 0.75>R>0.25, separated but overlapping; R<0.25, barely separated). Clinically-relevant ARGs were selected from a manually curated list prioritizing human health impacts of ARGs carried by infectious bacteria (**Supplementary Table S3**). Discriminatory ARGs were identified via the ExtrARG[16] machine learning based approach. ExtrARG is based on the extremely randomized trees classifier that uses a Bayesian strategy to optimize the parameters of the classifier and identify discriminatory ARGs based on the user-defined categorizing scheme or groups. LEfSe[72] enables biomarker discovery among class conditions within metagenomic samples. It first performs a non-parametric Kruskal-Wallis sum-rank test to detect features with differential abundance among the conditions of interest; it then applies an unpaired Wilcoxon rank-sum test to test for biological consistency. LEfSe uses linear discriminant analysis to estimate the effect size of each differentially abundant feature. Reads were assembled *de novo* in MetaStorm according to default parameters and scaffolds were mapped against the CARD and ACLAME databases. Network visualization was conducted using Gephi (version 0.8.2). Potential pathogenic ARG hosts were determined using the CARD prevalence tool, which examines the occurrence of each ARG of interest on NCBI archived pathogen chromosomes and assembled whole genome sequences, as well as plasmids.[37]

### Antibiotic analysis

Sample processing and analysis for antibiotics was conducted as previously described.[14] Solid phase extraction (SPE) was performed on 1 L filtered wastewater samples by conditioning Oasis HLB cartridges with acetonitrile and nanopure water before the water samples were loaded at a rate of 3-5 mL/min. Cartridges were dried under vacuum and then shipped to the University at Buffalo for elution and liquid chromatography with tandem mass spectrometry (LC-MS/MS) analysis. LC-MS/MS analysis was carried out using an Agilent 1200 LC system (Palo Alto, CA).

### Antibiotic loading

Anticipated antibiotic loads in sewage were calculated using the following expression:

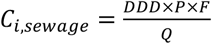

where C_*i,sewage*_(g/L) is the expected concentration of compound *i* in sewage, DDD (g/day) is the defined daily dose per person, P is the estimated sewershed population, F is the fraction of antibiotic excreted, and Q is the reported wastewater flow. Sewershed populations are notoriously challenging to obtain; however, they may be estimated based upon known excretion rates for widely used PPCPs such as carbamazepine, caffeine, and iopromide. For those systems where we detected one or more of these PPCPs, we estimated the sewershed population using:

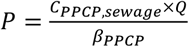

where C_*PPCP,sewage*_ is the measured concentration of carbamazepine (CBZ), caffeine (CAF), or iopromide (IOP) and β_*PPCP*_ is an empirical parameter relating sewage PPCP concentration to population. β_*CBZ*_ = 8.6 (± 1.4 at 95% confidence level) × 10^−8^ kg/day-person; β_*CAF*_ = 6.8 (± 3.2 at 95% confidence level) × 10^−7^ kg/day-person; β_*IOP*_ = 1.1 (± 0.19 at 95% confidence level) × 10^−5^kg/day-person. To date, β_*PPCP*_ values have only been reported for samples collected within Australia and thus this calculation inherently assumes it is universally applicable across all samples. Such an assumption requires future verification.

## Data Availability

The wastewater influent metagenomic datasets have been deposited in NCBI Short Read Archive (SRA) under bioproject PRJNA527877.

## Supporting Information

**SI:** Supplementary Figures (S1-S11) and Tables (S1-S3).

**Supplementary Data 1**: 16S Normalized Gene Counts

**Supplementary Data 2**: Clinically Relevant ARGs

**Supplementary Data 3**: Common ARGs

**Supplementary Data 4**: Discriminatory ARGs

**Supplementary Data 5**: Cross Resistance Genes

**Supplementary Data 6**: Antibiotic correlations

**Supplementary Data 7**: Manually curated database of ARGs included in analyses.

