## Supplemental Figures and Tables for "Wastewater Based Epidemiology Enabled Surveillance of Antibiotic Resistance"

**Supplementary Table S1.** Raw read and post-quality control (QC) sequence metrics. The average read length was 101 nucleotides. Quality control was performed via Trimmomatic.

| Sample_ID | Country | Visit_No | WWTP_No | RawReadPairs | ReadPairsAfterQC | %ReadsPairsPassingQC |
|---|---|---|---|---|---|---|
| IND1-P1-IN1 | IND | 1 | P1 | 13520740 | 13045504 | 96.49 |
| PHL1-P2-IN1 | PHL | 1 | P2 | 14919657 | 14332977 | 96.07 |
| USA1-P2-IN1 | USA | 1 | P2 | 13944727 | 13460770 | 96.53 |
| IND1-P1-IN2 | IND | 1 | P1 | 12303151 | 12093854 | 98.30 |
| CHE1-P2-IN1 | CHE | 1 | P2 | 15577543 | 15314202 | 98.31 |
| HKG1-P2-IN1 | HKG | 1 | P2 | 13401638 | 13174430 | 98.30 |
| PHL1-P1-IN1 | PHL | 1 | P1 | 12416972 | 12222754 | 98.44 |
| HKG2-P2-IN1 | HKG | 2 | P2 | 13932375 | 13707015 | 98.38 |
| IND1-P1-IN3 | IND | 1 | P1 | 11285980 | 11079243 | 98.17 |
| SWE1-P1-IN1 | SWE | 1 | P1 | 11973421 | 11801763 | 98.57 |
| USA1-P1-IN1 | USA | 1 | P1 | 13534261 | 13310751 | 98.35 |
| HKG2-P1-IN1 | HKG | 2 | P1 | 10069772 | 9883170 | 98.15 |
| IND1-P2-IN1 | IND | 1 | P2 | 14700590 | 14414214 | 98.05 |
| SWE1-P2-IN1 | SWE | 1 | P2 | 14784997 | 14524181 | 98.24 |
| USA1-P1-IN2 | USA | 1 | P1 | 13332573 | 13079484 | 98.10 |
| CHE1-P1-IN1 | CHE | 1 | P1 | 14454216 | 13932488 | 96.39 |

**Supplementary Table S2.** Top 25 most abundant ARGs for each WWTP

| India | | Hong Kong | | | | Philippines | | United States | | Switzerland | | Sweden | |
|---|---|---|---|---|---|---|---|---|---|---|---|---|---|
| **IND1-P1** | **IND1-P2** | **HKG1-P1** | **HKG1-P2** | **HKG2-P1** | **HKG2-P2** | **PHL1-P1** | **PHL1-P2** | **USA1-P1** | **USA1-P2** | **CHE1-P1** | **CHE1-P2** | **SWE1-P1** | **SWE1-P2** |
| sul1 | sul1 | QnrS2 | pmrE | dfrE | tetQ | pmrE | pmrE | msrE | pmrE | msrE | mphD | dfrE | dfrE |
| tetQ | pmrE | pmrE | dfrE | QnrS2 | pmrE | CfxA6 | CRP | mphD | dfrE | mphD | msrE | pmrE | pmrE |
| pmrE | tetQ | CRP | mefC | NPS-1 | dfrE | mel | tetQ | pmrE | mphD | dfrE | dfrE | mel | tetQ |
| ErmF | VEB-7 | dfrE | sul1 | pmrE | sul1 | tetQ | QnrS2 | dfrE | msrE | adeJ | pmrE | ErmB | adeJ |
| VEB-7 | tet(C) | QnrVC5 | mphG | CRP | ErmF | dfrE | sul1 | tet(39) | tetQ | adeK | adeJ | mefA | msrE |
| tet(C) | ErmF | msbA | tetQ | msbA | CRP | sul1 | CfxA6 | adeJ | CRP | pmrE | tetQ | tetQ | mphD |
| CfxA4 | dfrE | APH(6)-Id | CRP | Ec-CpxR | ErmB | mefA | tet(C) | CRP | qacH | abeM | ErmB | adeJ | abeM |
| CRP | CfxA4 | NPS-1 | tetX | ErmB | CfxA6 | tet37 | AAC(6')-Ib7 | tetQ | sul1 | adeI | adeK | lnuD | adeK |
| CfxA6 | CRP | Ec-CpxR | ErmF | APH(3")-Ib | mefC | CRP | vgaC | sul1 | ErmF | tetQ | ANT(3")-IIc | abeM | ErmB |
| dfrE | AAC(6')-Ib7 | APH(3")-Ib | ErmB | macB | mphG | lsaE | qacH | macB | OXA-16 | tet(39) | abeM | adeK | tet(39) |
| APH(6)-Id | APH(3")-Ib | emrR | tet(C) | tetQ | AAC(6')-IId | APH(6)-Id | Eci-CpxR | msbA | tet(C) | mexK | adeI | macB | adeI |
| msrE | mel | macB | AAC(6')-IId | mtrA | CfxA4 | qacH | dfrE | adeK | OXA-210 | qacH | qacH | tetW | mefA |
| mphD | mefA | sul1 | qacH | APH(6)-Id | mel | tet(C) | QnrVC5 | qacH | tet(39) | ANT(3")-IIc | mexK | tet(39) | macB |
| mel | qacH | tet34 | AAC(6')-Ib7 | tet34 | MCR-3 | APH(3")-Ib | msbA | mexK | mefA | macB | CRP | adeI | mel |
| mefA | APH(6)-Id | mel | VEB-7 | tet32 | bacA | AAC(6')-Ib7 | QnrVC3 | mphG | macB | tetW | macB | mexK | mexK |
| AAC(6')-Ib7 | msrE | bacA | EreA | msrB | macB | macB | msrE | abeM | tetW | adeN | tet(39) | CfxA6 | tetW |
| APH(3")-Ib | cmlA5 | acrB | msbA | lnuC | VEB-7 | lnuD | mphD | mefC | QnrS2 | mphG | Eci-CpxR | qacH | Ec-CpxR |
| cmlA5 | aadA22 | tet(35) | aadA8 | sul1 | msbA | lnuB | patA | OXA-16 | CfxA4 | CfxA6 | msbA | efrB | msbA |
| OXA-16 | mphD | lnuC | mel | bacA | mefA | ErmF | Ec-LamB | ErmF | msbA | ErmB | ErmF | lmrD | CRP |
| tetW | tet36 | acrD | APH(3")-Ib | QnrS4 | AAC(6')-Ib7 | catQ | pmrF | adeI | AAC(6')-Ib7 | mefC | CfxA6 | CRP | adeN |
| catQ | tetW | vgaC | mefA | tetW | aadA8 | ErmB | cpxA | bacA | mel | OXA-333 | tetW | tet37 | qacH |
| arr-2 | arr-2 | QnrS4 | APH(6)-Id | kdpE | tetW | CfxA4 | bacA | tet(C) | AER-1 | OXA-334 | bacA | ErmF | CfxA6 |
| aadA22 | CfxA6 | ErmB | macB | emrR | tetO | aadA8 | tet(39) | Ec-CpxR | ErmB | ErmF | mphG | msbA | CfxA4 |
| qacH | tetM | QnrS5 | Escherichia-coli-CpxR | adeJ | Escherichia-coli-CpxR | msbA | acrB | OXA-210 | OXA-74 | msbA | APH(3")-Ib | msrE | ANT(3")-IIc |
| tet36 | macB | tetQ | mefB | acrB | tet32 | QnrS2 | APH(3")-Ib | MexB | acrB | APH(3")-Ib | APH(6)-Id | mphD | APH(3")-Ib |

**Supplementary Table S3**: Known pathogen hosts for discriminatory ARGs and presence of ARGs on plasmids in selected pathogens

| Gene | Known Pathogen Hosts (out of 82 important pathogens)[1] | Plasmid-borne in pathogens? |
|---|---|---|
| adeJ | *Acinetobacter baumannii, Acinetobacter nosocomialis, Escherichia coli, Helicobacter pylori, Pseudomonas aeruginosa, Staphylococcus aeureus* | No |
| adeK | *Acinetobacter baumannii, Acinetobacter nosocomialis, Klebsiella pneumoniae, Pseudomonas aeruginosa, Staphylococcus aureus* | No |
| abeM | *Acinetobacter baumannii, Acinetobacter nosocomialis, Escherichia coli, Klebsiella pneumoniae* | No |
| tet(39) | *Acinetobacter baumannii, Acinetobacter nosocomialis, Klebsiella oxytoca, Klebsiella pneumoniae* | Yes |
| adeI | *Acinetobacter baumannii, Acinetobacter nosocomialis* | No |
| mexK | *Acinetobacter baumannii, Enterobacter cloacae, Klebsiella pneumonia, Pseudomonas fluorescens, Pseudomonas putida* | No |
| adeN | *Acinetobacter baumannii, Acinetobacter nosocomialis, Klebsiella pneumonia* | No |
| OXA-333 | *Acinetobacter baumannii, Acinetobacter nosocomialis* | No |
| OXA-334 | NA | |
| OXA-211 | *Acinetobacter nosocomialis* | No |
| MuxB | *Acinetobacter baumannii, Enterobacter cloacae, Klebsiella pneumoniae, Pseudomonas aeruginosa, Salmonella enterica* | No |
| OXA-363 | *Acinetobacter lwoffii, Acinetobacter nosocomialis* | No |
| MOX-9 | NA | No |
| CpxR | *Escherichia coli, Salmonella* spp. | No |
| mtrD | *Neisseria gonorrhoeae, Neisseria meningitidis* | No |
| mdsB | *Salmonella enterica* | No |
| MCR-2 | *Shigella sonnei* | No |
| adeG | *Acinetobacter baumannii, Acinetobacter nosocomialis, Clostridioides difficile, Klebsiella pneumonia, Salmonella enterica* | No |
| FOX-1 | NA | No |
| mexQ | *Acinetobacter baumannii, Enterobacter cloacae, Klebsiella pneumoniae, Pseudomonas aeruginosa* | No |
| OprM | *Acinetobacter baumannii, Enterobacter cloacae, Klebsiella pneumoniae, Pseudomonas aeruginosa* | No |
| PmpM | NA | |
| axyY | *Achromobacter xylosoxidians, Escherichia coli* | No |
| cmlA1 | *Acinetobacter baumannii, Escherichia coli, Klebsiella pneumoniae, Proteus mirabilis, Pseudomonas aeruginosa, Pseudmonoas putida, Pseudomonas stutzeri, Salmonella eterica, Shigella flexneri* | Yes |
| sul3 | *Citrobacter freundii, Escherichia coli, Klebsiella aerogenes, Klebsiella pneumoniae, Proteus mirabilis, Salmonella enterica, Shigella flexneri, Shigella sonnei, Yersinia enterocolitica* | Yes |
| CARB-4 | *Acinetobacter baumannii, Pseudomonas putida, Vibrio cholerae* | Yes |
| evgS | *Citrobacter freundii, Enterobacter cloacae, Enterobacter kobei, Enterococcus faecium, Escherichia coli, Klebsiella oxytoca, Klebsiella* | Yes |

| | | |
|---|---|---|
| | *pneumoniae, Pseudomonas aeruginosa, Salmonella enterica, Serratia marcescens, Shigella dysenteriae, Shigella flexneri, Shigella sonnei, Staphyloccus aureus, Staphyloccus epidermidis* | |
| dfrA5 | *Acinetobacter baumannii, Enterobacter cloacae, Escherichia coli, Klebsiella oxytoca, Klebsiella pneumoniae, Proteus vulgaris, Pseudomonas aeruginosa, Salmonella enterica, Serratia marcescens, Shigella dysenteriae, Shigella flexneri, Shigella sonnei* | Yes |
| CARB-2 | *Salmonella enterica* | No |
| AAC(6')-Ib11 | *Enterobacter asburiae, Escherichia coli, Klebsiella pneumoniae, Pseudomonas aeruginosa, Pseudomonas stutzeri, Serratia marcescens* | No |
| AAC(6')-30/AAC(6')-Ib' | *Acinetobacter baumannii, Escherichia coli, Klebsiella aerogenes, Klebsiella pneumoniae, Pseudomonas aeruginosa* | Yes |
| pp-flo | *Escherichia coli, Klebsiella pneumoniae, Psedomonas aeruginosa, Salmonella enterica* | No |
| ErmT | *Enterococcus faecium, Escherichia coli, Staphyloccus aureus, Streptococcus agalactiae, Streptococcus pyogenes* | Yes |
| catB8 | *Acinetobacter baumannii, Enterobacter asburiae, Enterobacter cloacae, Klebsiella oxytoca, Klebsiella pneumoniae, Providencia rettgeri, Pseudomonas aeruginosa, Raoultella planticola, Shigella sonnei* | Yes |
| aadA2 | *Acinetobacter baumannii, Acinetobacter nosocomialis, Citrobacter freundii, Citrobacter koseri, Enterobacter asburiae, Enterobacter cloacae, Enterobacter hormaechei, Enterobacter kobei, Escherichia coli, Klebsiella aerogenes, Klebsiella oxytoca, Klebsiella pneumoniae, Morganella morganii, Proteus mirabilis, Providencia rettgeri, Pseudomonas aeruginosa, Raoultella planticola, Salmonella enterica, Shigella dysenteriae, Shigella flexneri, Shigella sonnei, Stenotrophomonas maltophilia, Vibrio parahaemolyticus, Yersinia enterocolitica, Yersinia pestis* | Yes |
| QnrVC6 | *Pseudomonas aeruginosa, Pseudomonas putida* | Yes |
| cmlA4 | *Citrobacter frendii, Escherichia coli, Klebsiella aerogenes, Klebsiella pneumoniae, Pseudomonas aeruginosa, Salmonella enterica* | Yes |
| dfrA1 | *Acinetobacter baumannii, Citrobacter freundii, Enterobacter asburiae, Enterobacter cloacae, Enterobacter hormaechei, Enterobacter kobei, Escherichia coli, Klebsiella aerogenes, Klebsiella oxytoca, Klebsiella pneumoniae, Morganella morganii, Proteus mirabilis, Proteus penneri, Proteus vulgaris, Providencia rettgeri, Providencia stuartii, Pseudomonas aeruginosa, Pseudomonas stutzeri, Raoultella planticola, Salmonella enterica, Serratia marcescens, Shigella dysenteriae, Shigella flexneri, Shigella sonnei, Vibrio cholerae, Vibrio parahaemolyticus, Vibrio vulnificus, Yersinia enterocolitica* | Yes |
| sul2 | *Achromobacter xylosoxidans, Acinetobacter baumannii, Acinetobacter lwoffii, Acinetobacter nosocomialis, Citrobacter amalonaticus, Citrobacter freundii, Citrobacter koseri, Enterobacter asburiae, Enterobacter cloacae, Enterobacter hormaechei, Enterobacter kobei, Escherichia coli, Klebsiella aerogenes, Klebsiella oxytoca, Klebsiella pneumoniae, Morganella morganii, Proteus mirabilis, Proteus vulgaris, Providencia rettgeri, Providencia stuartii, Pseudomonas aeruginosa, Pseudomonas stutzeri, Salmonella enterica, Serratia marcescens, Shigella dysenteriae, Shigella* | Yes |

| | | |
|---|---|---|
| | *flexneri, Shigella sonnei, Stenotrophomonas maltophilia, Vibrio cholerae, Vibrio parahaemolyticus, Vibrio vulnificus, Yersinia enterocolitica, Yersinia pestis* | |
| QnrVC3 | NA | |
| catB3 | *Acinetobacter baumannii, Citrobacter freundii, Enterobacter asburiae, Enterobacter cloacae, Enterobacter hormaechei, Enterobacter kobei, Escherichia coli, Klebsiella aerogenes, Klebsiella oxytoca, Klebsiella pneumoniae, Morganella morganii, Proteus mirabilis, Providencia rettgeri, Pseudomonas aeruginosa, Raoultella planticola, Salmonella enterica, Shigella flexneri* | Yes |
| arr-2 | *Acinetobacter baumannii, Citrobacter freundii, Enterobacter cloacae, Enterobacter hormaechei, Escherichia coli, Klebsiella oxytoca, Klebsiella pneumoniae, Proteus mirabilis, Providencia rettgeri, Providencia stuartii, Pseudomonas aeruginosa, Pseudomonas stutzeri, Serratia marcescens* | Yes |
| EreA | *Escherichia coli, Klebsiella oxytoca, Proteus mirabilis, Salmonella enterica* | Yes |
| aadA22 | *Escherichia coli, Klebsiella pneumoniae, Salmonella entericap, Shigella sonnei* | Yes |
| aadA8 | *Acinetobacter baumannii, Citrobacter freundii, Escherichia coli, Klebsiella aerogenes, Klebsiella oxytoca, Klebsiella pneumoniae, Providencia rettgeri, Pseudomonas aeruginosa, Salmonella enterica, Serratia marcescens, Shigella flexneri, Shigella sonnei, Yersinia enterocolitica* | No |
| AAC(6')-Iid | NA | |
| VEB-7 | *Acinetobacter baumannii* | No |
| sul1 | *Achromobacter xylosoxidans, Acinetobacter baumannii, Acinetobacter nosocomialis, Burkholderia cenocepacia, Citrobacter freundii, Citrobacter koseri, Enterobacter asburiae, Enterobacter cloacae, Enterobacter hormaechei, Enterobacter kobei, Escherichia coli, Klebsiella aerogenes, Klebsiella oxytoca, Klebsiella pneumoniae, Morganella morganii, Proteus mirabilis, Providencia rettgeri, Providencia stuartii, Pseudomonas aeruginosa, Pseudomonas putida, Pseudomonas stutzeri, Raoultella planticola, Salmonella enterica, Serratia marcescens, Shigella dysenteriae, Shigella flexneri, Shigella sonnei, Stenotrophomonas maltophilia, Vibrio cholerae, Vibrio parahaemolyticus, Vibrio vulnificus, Yersinia enterocolitica, Yersinia pestis* | Yes |

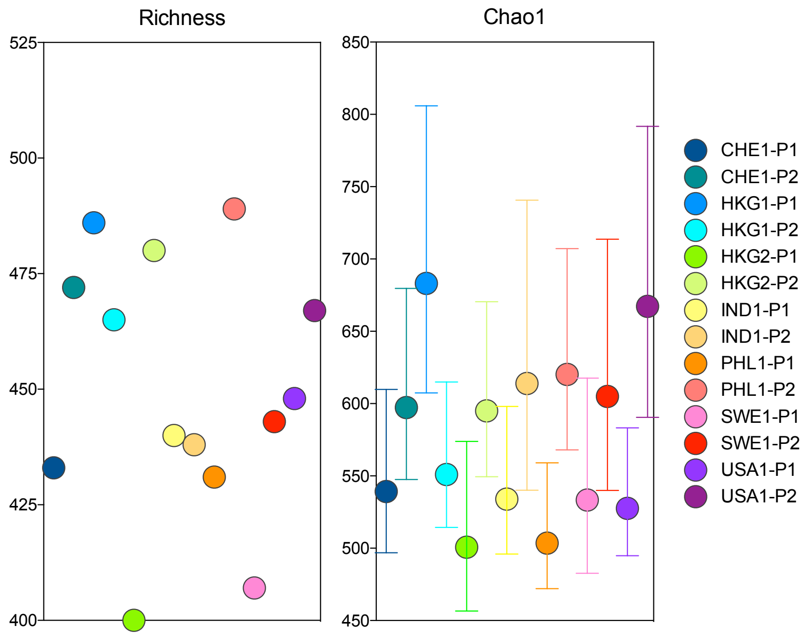


**Supplementary Figure S1:** Observed number of unique ARGs (Richness) and Chao1 index based on MetaStorm annotations to CARD version 1.2.1 database. Error bars represent the Chao1 95% upper and lower confidence intervals. Sample names refer to countries (IND: India, PHL: Philippines, USA: United States, CHE: Switzerland, HKG: Hong Kong, SWE: Sweden) along with the visit number and WWTP plant (P) number.

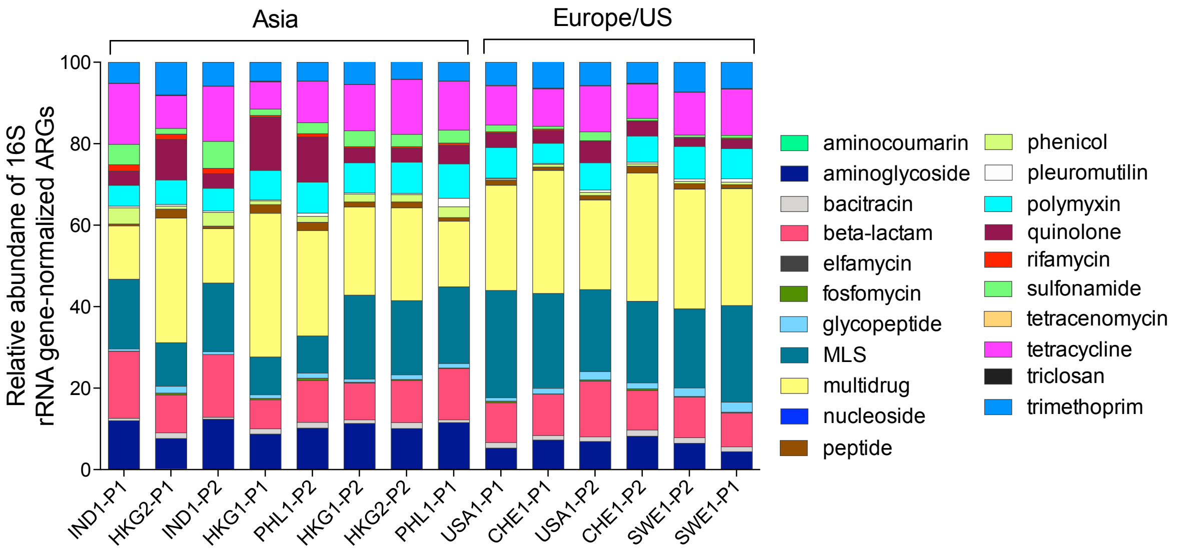


**Supplementary Figure S2.** Percent relative abundance of ARG classes detected across the sewage samples. ARGs were annotated via MetaStorm using CARD version 1.2.1. ARG categories were assigned in-house according to the categories specified in Supplementary Data 1. Genes corresponding to two or more categories were labeled as “multidrug.” ARG abundances were normalized via MetaStorm to 16S rRNA gene abundances. Sample names refer to countries (IND: India, PHL: Philippines, USA: United States, CHE: Switzerland, HKG: Hong Kong, SWE: Sweden) along with the visit number and WWTP plant (P) number.

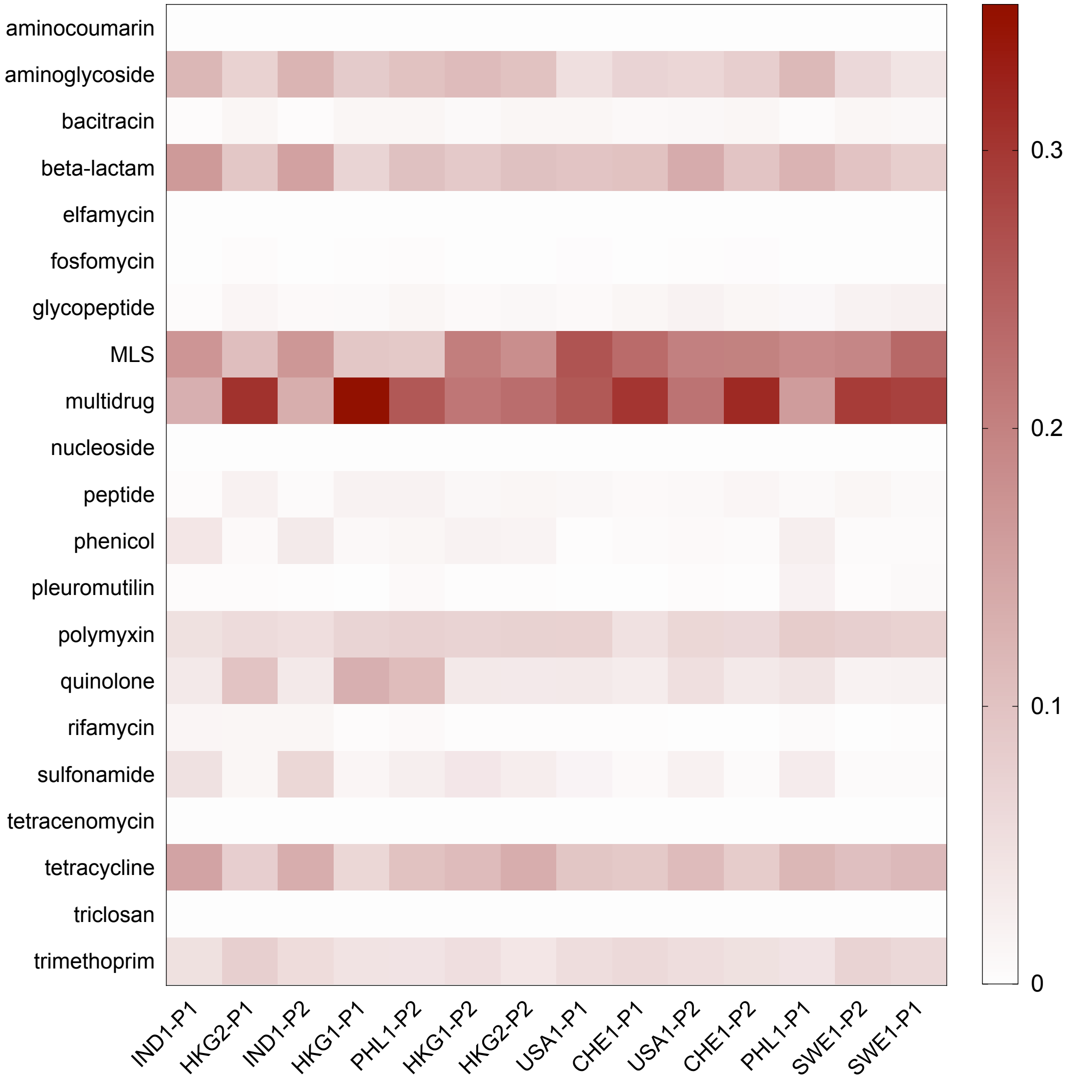


**Supplementary Figure S3.** Heatmap visualization of ARG relative abundance by category. ARGs were annotated via MetaStorm using CARD version 1.2.1. ARG categories were assigned in-house according to the categories specified in Supplementary Data 1. Genes corresponding to two or more categories were labeled as “multidrug.” ARG abundances were normalized via MetaStorm to 16S rRNA gene abundances. Regulatory and housekeeping genes are not shown. Sample names refer to countries (IND: India, PHL: Philippines, USA: United States, CHE: Switzerland, HKG: Hong Kong, SWE: Sweden) along with the visit number and WWTP plant (P) number.

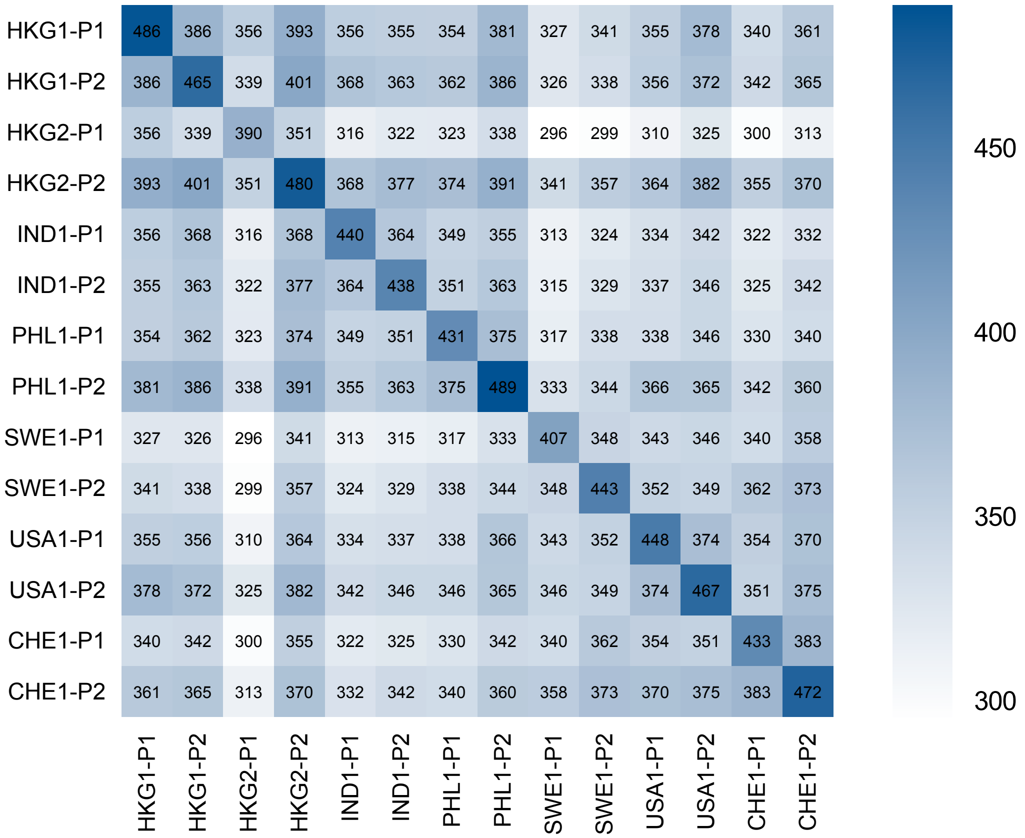


**Supplementary Figure S4.** Number of shared annotated ARGs between each site. Diagonal represents the number of observed ARGs at each site (i.e., the ARG alpha diversity).

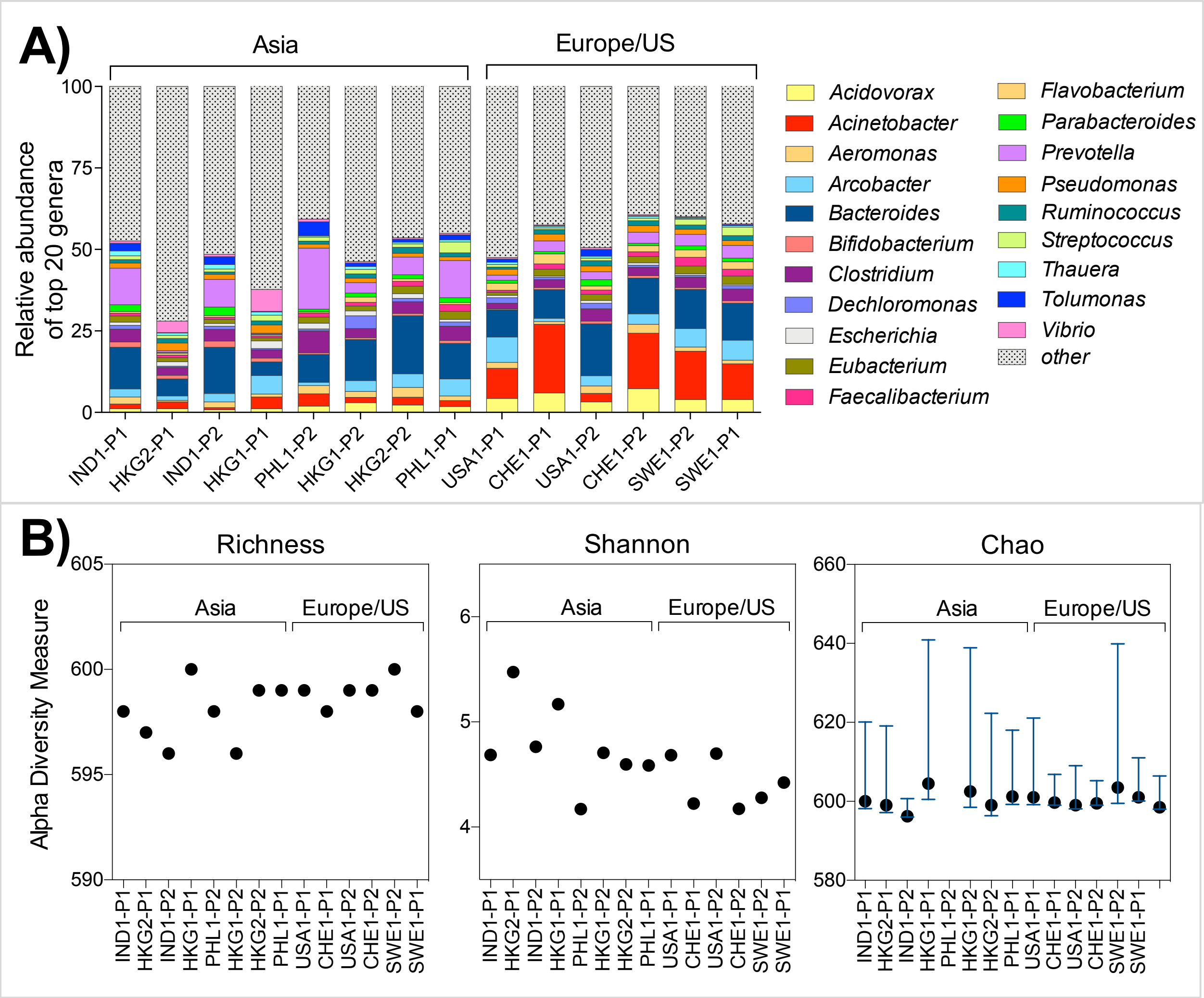


**Supplementary Figure S5**: Bacterial genera in sewage samples. (A) Relative abundance of top 20 bacterial genera in sewage. Genus-level annotations were done via MG-RAST using the Refseq database. (B) Alpha diversity measures of WWTPs with respect to bacterial genera. Richness, Shannon, and Chao metrics. Error bars represent the Chao1 95% upper and lower confidence intervals. Sample names refer to countries (IND: India, PHL: Philippines, USA: United States, CHE: Switzerland, HKG: Hong Kong, SWE: Sweden) along with the visit number and WWTP plant (P) number.

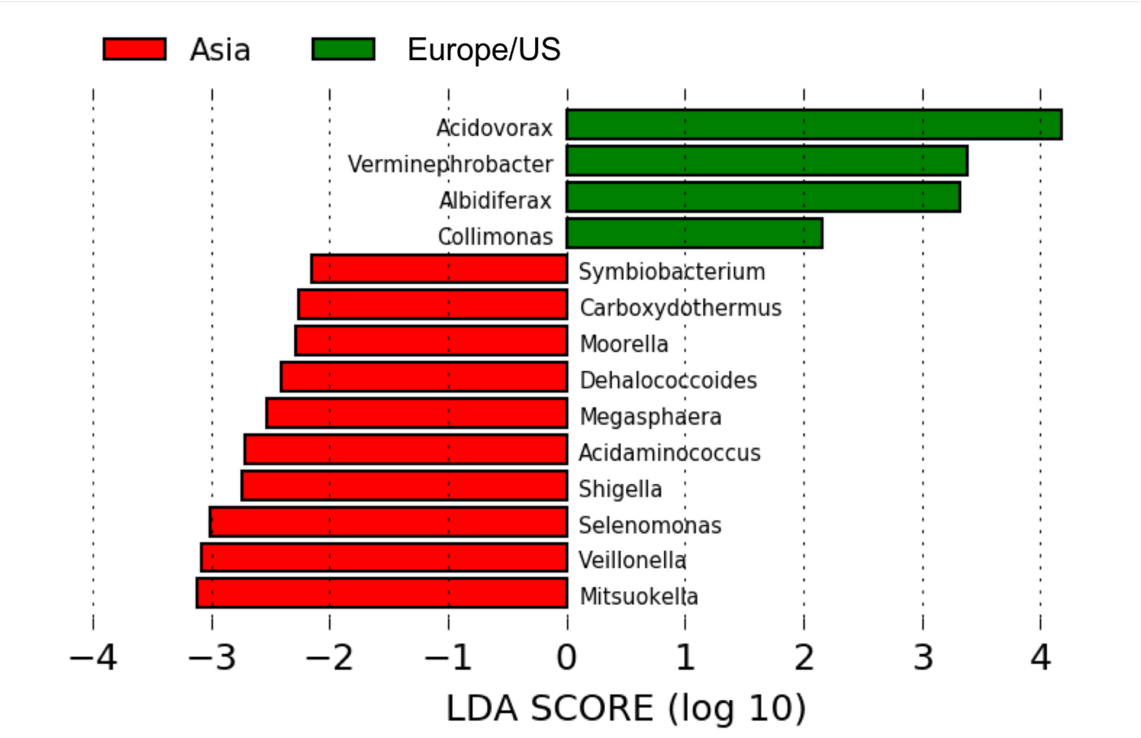


**Supplementary Figure S6**. Lefse-identified classifying genera for Asia and Europe/America regions.

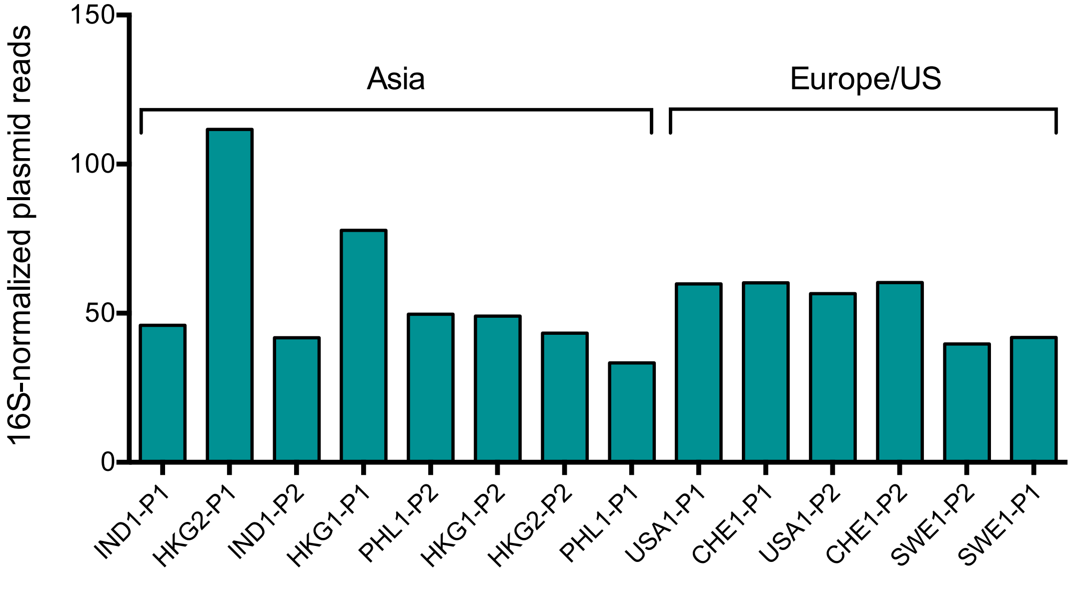


**Supplementary Figure S7**: Total relative abundance of plasmid-associated genes in WWTP influents, annotated via MetaStorm using the ACLAME database version 0.4. Plasmid-associated gene abundances were normalized to 16S rRNA gene abundances. Samples refer to countries (IND: India, PHL: Philippines, USA: United States, CHE: Switzerland, HKG: Hong Kong, SWE: Sweden) along with the visit number and WWTP plant.

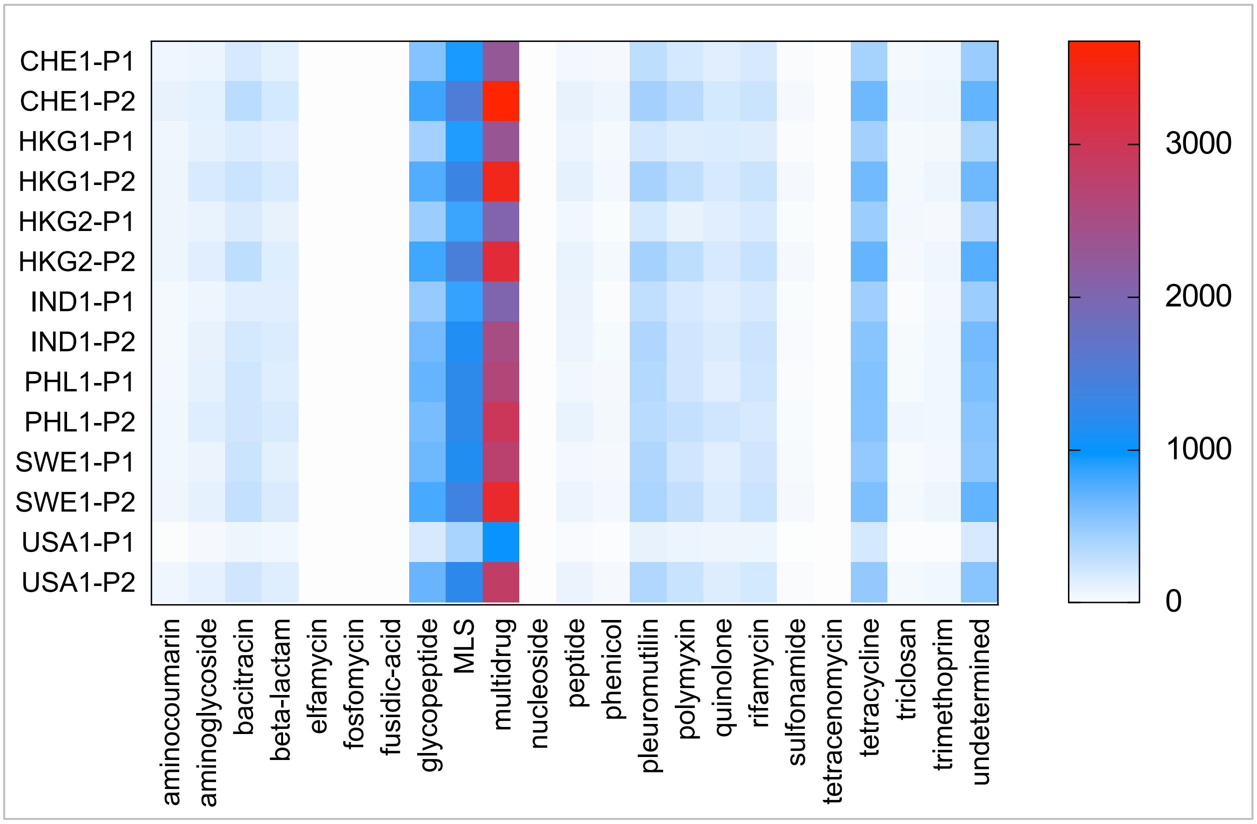


**Supplementary Figure S8**. Counts of ARGs conferring resistance to the specified antibiotic categories within the assembled scaffolds. Color scale represents the number of times that each ARG category is found in the scaffolds.

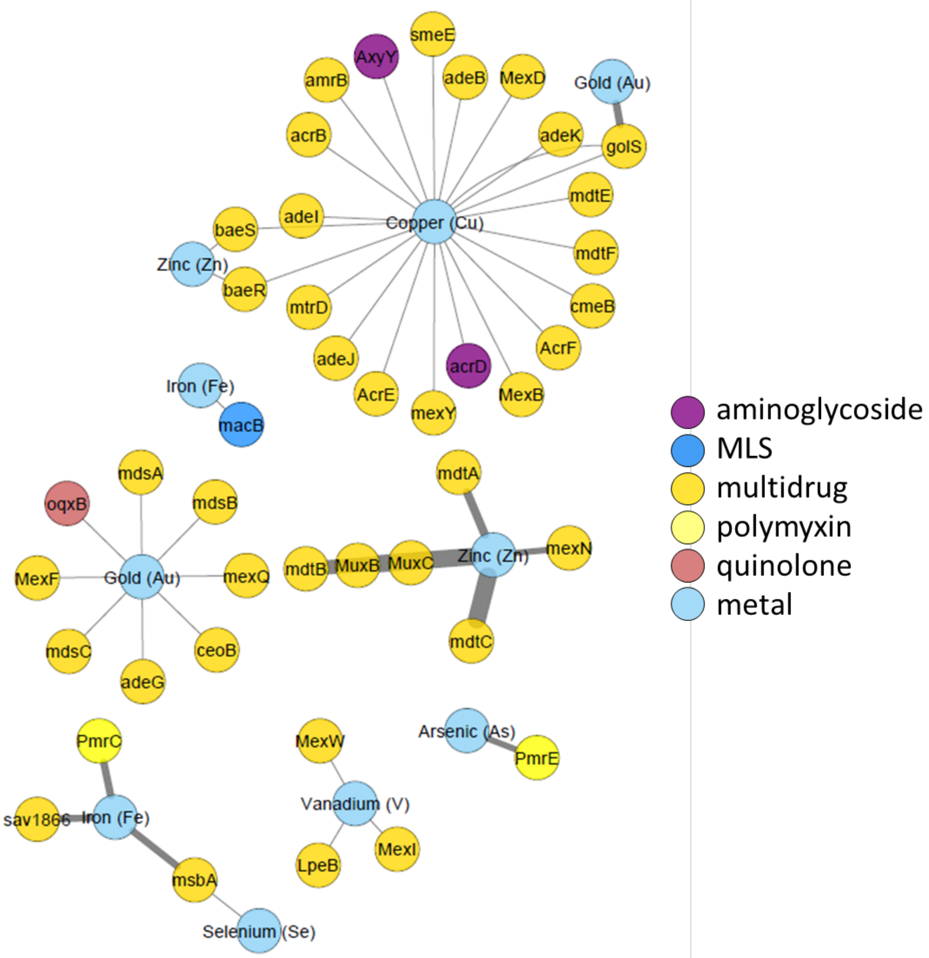


**Supplementary Figure S9**: Co-occurrence of ARGs and MRGs on *de novo* assembled scaffolds generated by shotgun metagenomic sequencing reads pooled from all samples. Proximity of nodes and width of lines indicate frequency of associations between genes. Node diameter is proportional to the number of co-occurrences for that gene. Co-occurences with fewer than 3 instances were excluded from the network analysis rendering. MLS = macrolide-lincosamide-streptogramin resistance.

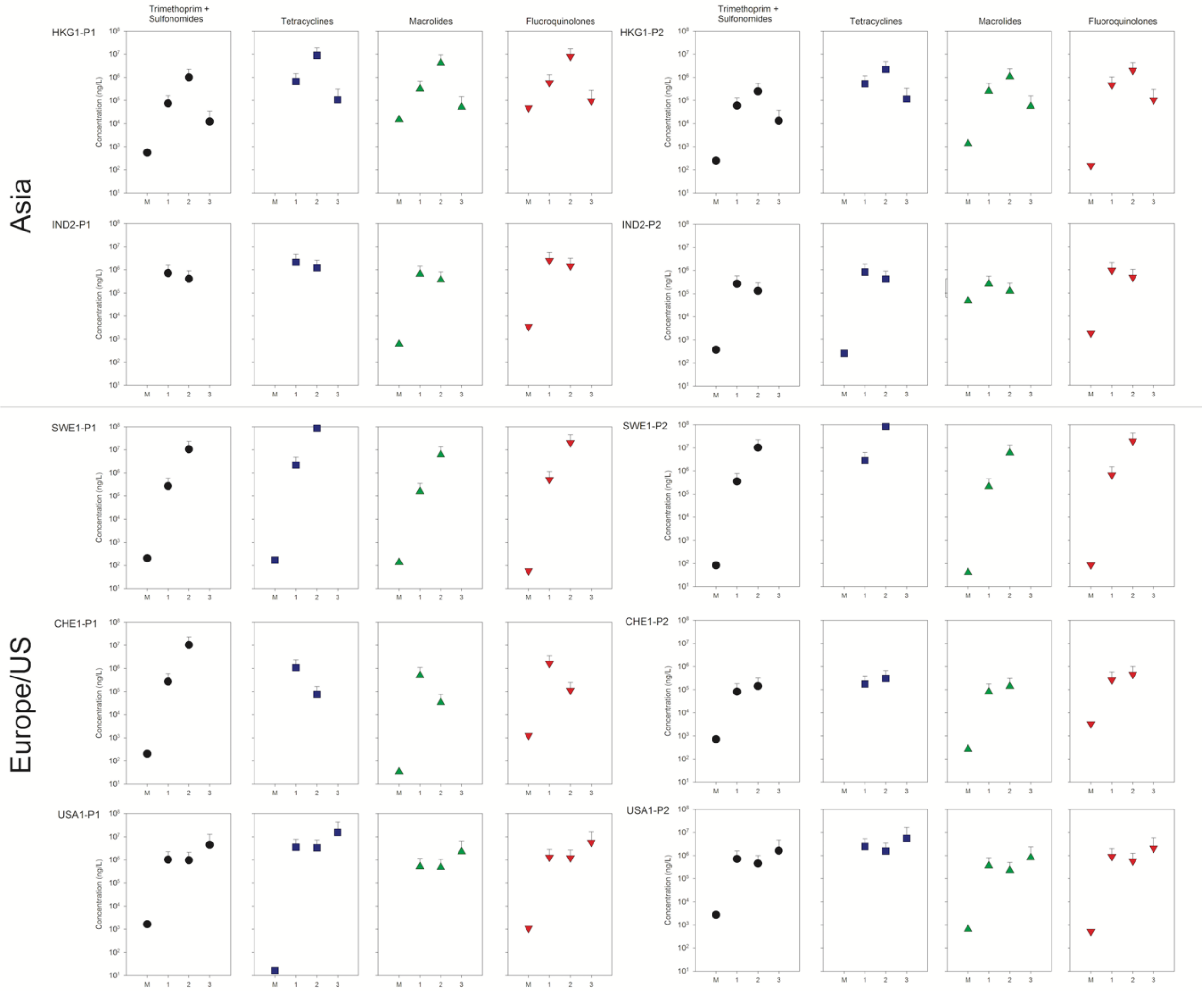


**Supplementary Figure S10**. Measured antibiotic and theoretical concentrations in influent sewage. Nomenclature: M – measured concentrations, 1,2,3 – concentrations predicted based upon measured caffeine (1), carbamazepine (2), and iopromide (3) concentrations. Sample names refer to countries (IND: India, PHL: Philippines, USA: United States, CHE: Switzerland, HKG: Hong Kong, SWE: Sweden) along with the visit number and WWTP plant (P) number.

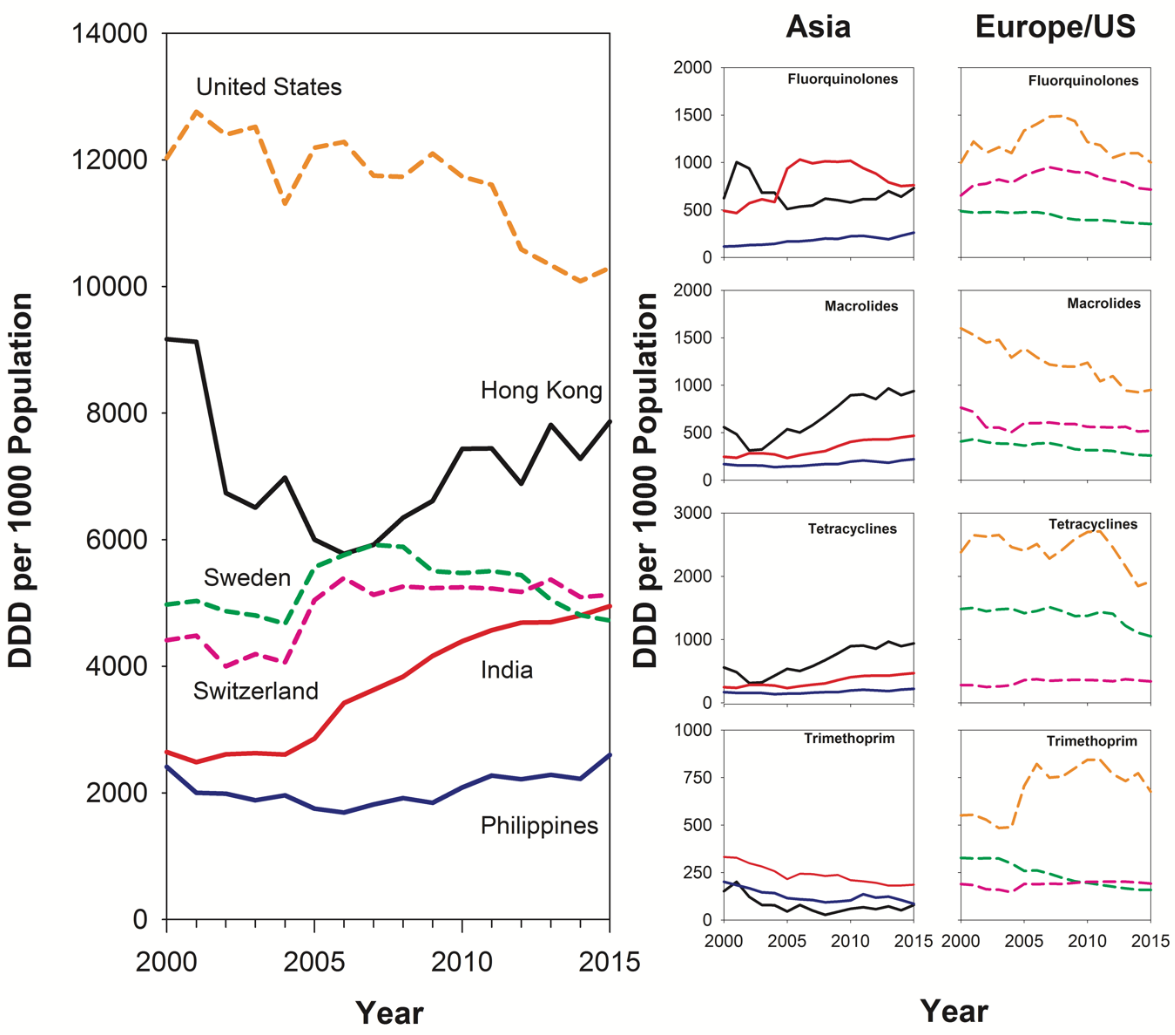


**Supplementary Figure S11**. Estimated antibiotic consumption rates for Asia (Hong Kong, India, Philippines) and Europe/US (Sweden, Switzerland, United States) in defined daily doses (DDDs) per 1000 inhabitants. The left panel reflects total antibiotic usage within the indicated location. The right panels provide detailed trends for specific drugs and drug classes. The color and line weighting scheme in the left panel is used throughout the Figure. Data source: https://resistancemap.cddep.org
